# Early prediction of preeclampsia in pregnancy with circulating cell-free RNA

**DOI:** 10.1101/2021.03.11.21253393

**Authors:** Mira N. Moufarrej, Sevahn K. Vorperian, Ronald J. Wong, Ana A. Campos, Cecele C. Quaintance, Rene V. Sit, Michelle Tan, Angela M. Detweiler, Honey Mekonen, Norma F. Neff, Maurice L. Druzin, Virginia D. Winn, Gary M. Shaw, David K. Stevenson, Stephen R. Quake

## Abstract

Liquid biopsies that measure circulating cell-free RNA (cfRNA) offer an unprecedented opportunity to noninvasively study the development of pregnancy-related complications and to bridge gaps in clinical care. Here, we used 404 blood samples from 199 pregnant mothers to identify and validate cfRNA transcriptomic changes that are associated with preeclampsia (PE), a multi-organ syndrome which is the second largest cause of maternal death globally. We find that changes in cfRNA gene expression between normotensive (NT) and preeclamptic mothers are striking and stable early in gestation, well before the onset of symptoms. These changes are enriched for genes specific to neuromuscular, endothelial, and immune cell types and tissues that reflect important aspects of PE physiology and suggest new hypotheses for disease progression. This enabled identification and independent validation of a panel of 18 genes whose measurement between 5–16 weeks of gestation can form the basis of a liquid biopsy test that would identify mothers at risk of PE well before the clinical symptoms manifest themselves. Finally, we demonstrate that cfRNA changes reflect the multifactorial nature of PE and provide a means to non-invasively monitor maternal organ health. Tests based on these observations could help predict and manage who is at risk for PE, an important and until now unachieved objective for obstetric care.

## Introduction

Advances in obstetrics and neonatology have significantly mitigated many of the adverse pregnancy outcomes related to preterm birth (PTB) and preeclampsia (PE) ^1,2^. Nonetheless, the standards of care implemented today focus on how to treat a mother and child once a complication has been diagnosed, proving both insufficient and costly ^3–6^: PE and related hypertensive disorders cause 14% of maternal deaths each year globally, second only to hemorrhage ^7^, and cost nearly $2B in care in the first year following delivery ^5^. Worse, 3 out of 5 maternal deaths in the US are preventable and often associated with a missed or delayed complication diagnosis ^8^. Such outcomes highlight the need for novel tools that would aid in identifying which mothers are at risk for hypertensive diseases, such as PE, before clinical presentation. Indeed, early prediction, which has not been achieved to date, may enable prevention or reduction of a pregnant mother’s risk of developing PE ^9,10^ if coupled with appropriate treatment.

PE is a leading pregnancy-related complication that globally affects 4–5% of pregnancies ^11–13^ and is associated with a significant increase in adverse maternal and perinatal outcomes ^14–18^. Long-term, PE presents an increased maternal risk for cardiovascular ^19,20^ and kidney ^21,22^ diseases. Formally defined as new-onset hypertension with proteinuria or other organ damage (e.g., liver, brain) occurring after 20 weeks of gestation ^23^, PE can clinically manifest anytime thereafter, including into the post-partum period ^24^. Detection and diagnosis have proven challenging both because early signs such as headaches and nausea can be easily confused with generalized pregnancy discomfort and because PE shares many signs and symptoms with other common complications like gestational thrombocytopenia and chronic hypertension.

To date, no recommended test exists that can predict the future onset of PE early in pregnancy ^9^. Investigational approaches that measure biochemical signals including the measurement of two angiogenic factors (soluble fms-like tyrosine kinase-1 (sFlt1), placental growth factor (PlGF)) in the second and third trimesters ^25,26^ have at best only moderate positive predictive values (8–33%)^27^. A clinical test could guide the prophylactic use of low-dose aspirin, which has been shown to safely reduce the risk of PE if initially administered before 16 weeks of gestation ^10,28,29^. Liquid biopsies that measure plasma cell-free RNA (cfRNA) suggest a means to bridge this gap in clinical care; however, until recently, such work often failed to progress beyond initial discovery ^30^. Recent efforts have instead either focused on confirmation of PE at clinical diagnosis ^31,32^ or on limited individuals earlier in pregnancy with encouraging but unvalidated results ^33^. Consequently, the prediction of PE early in gestation, before symptoms present, when such a test would be most useful to guide the prophylactic use of potential therapeutics remains a key objective to improve obstetric care ^9^.

Clinical care may also be improved by a better understanding of PE’s pathogenesis. PE is which is specific to humans ^34^ and a few non-human primates ^35^, and consequently, elucidating its pathogenesis has proved challenging. Broadly, it is accepted that PE occurs in two stages – abnormal placentation early in pregnancy followed by systemic endothelial dysfunction ^15,34,36^. Because PE can clinically present any time after 20 weeks of gestation and with a diversity of symptoms, significant effort has been made to subclassify the disease based upon the timing of onset as a proxy for pathology ^37,38^; however, debate over the significance of such subtyping is ongoing ^34,36,39,40^. Separate efforts have focused on subtyping PE molecularly using placental gene expression and histology ^41,42^. Since cfRNA is derived from many tissues in the body ^43,44^, noninvasive liquid biopsies present a potential means to indirectly observe pathogenesis in real time and to identify physiological changes associated with PE for proposed subtypes both prior to and at diagnosis.

Here, we report that cfRNA transcriptomic changes can distinguish between normotensive (NT) and PE pregnancies throughout the course of pregnancy, irrespective of PE subtype. Interestingly, the majority of these cfRNA changes are most striking early in pregnancy – well before the onset of symptomns - suggesting that the identified cfRNA signal may correlate with PE pathogenesis, which is thought to also occur at this time. Indeed, gene ontology (GO) and tissue and cell type of origin analyses identified pathways, tissues, and cell types that both reflect known PE physiology and suggest new means to stratify the disease. These observations enabled us to identify and independently validate a panel of 18 genes that when measured between 5–16 weeks of gestation form a predictive signature of which mothers are at risk for developing PE. Finally, we demonstrate that cfRNA signals reflect the multifactorial nature of PE and provide a means to noninvasively monitor maternal organ health. Taken together, these results show that cfRNA measurements can form the basis for clinically relevant tests that would predict PE months before presentation, manage who is at risk for specific organ damage, and help characterize the pathogenesis of PE in real time.

## Results

### Clinical study design

To identify changes associated with PE well before traditional diagnosis, we designed a prospective study and recruited pregnant mothers at their first clinical visit to Stanford’s Lucile Packard Children’s Hospital, between 5–12 weeks of gestation, of which 131 were included in this study (94 NT, 37 with PE). For each participant, we analyzed cfRNA for samples collected before or at 12 weeks, between 13-20 weeks, and at or after 23 weeks of gestation, and post-partum (0–4 weeks after delivery). We then split this larger group into Discovery (n = 88, [60 NT, 28 with PE]) and Validation 1 (n = 43, [34 NT, 9 with PE]) cohorts. We also obtained samples from an independent cohort collected at several separate institutions (Validation 2), which consisted of 89 samples collected prior to 16 weeks of gestation from 87 mothers (61 NT, 26 with PE) (Fig 1A).

**Figure 1.**
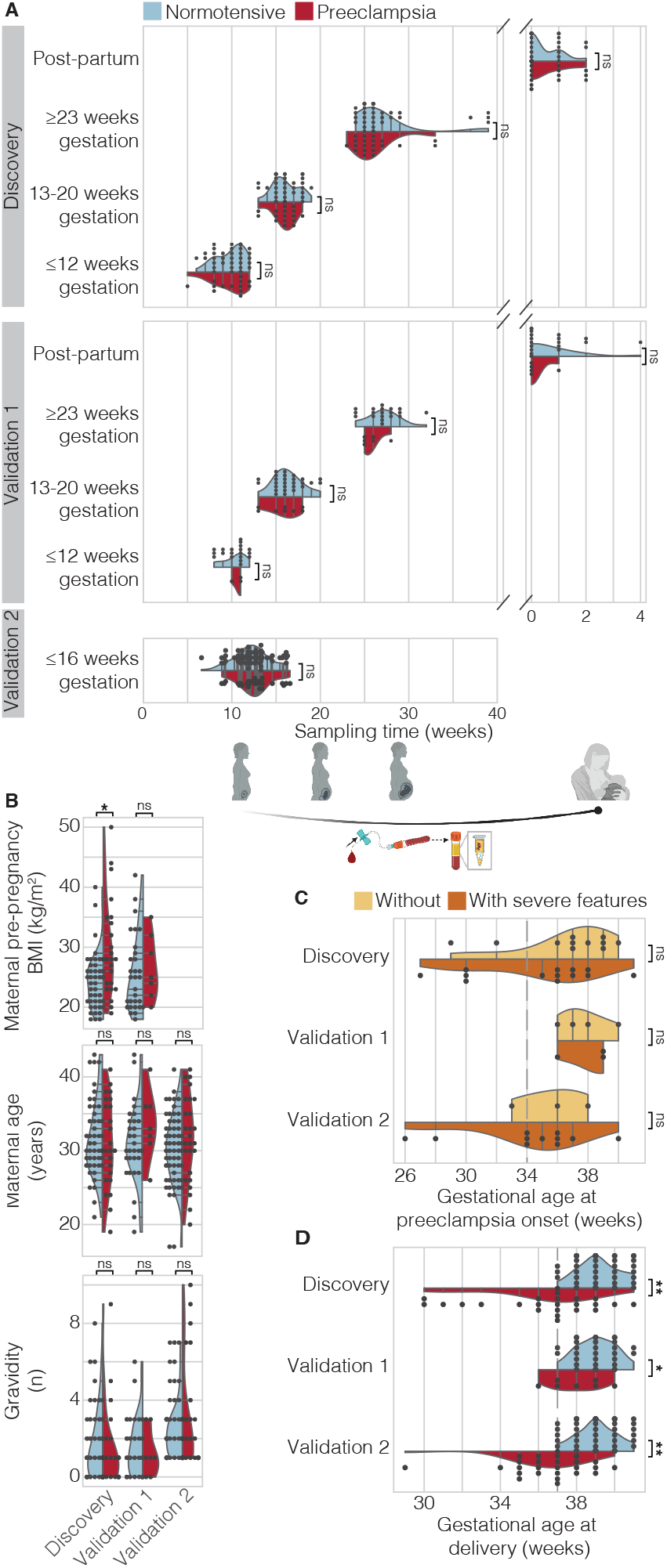
Comparing sample, maternal, and pregnancy characteristics for NT and PE groups across all cohorts. Panels illustrate matched sample collection time (weeks) in (**A**), maternal characteristics (Top to bottom: BMI, age, and gravidity) in (**B**), matched gestational age at PE onset regardless of PE symptom severity in (**C**), and gestational age at delivery in (**D**) for Discovery, Validation 1, and Validation 2 cohorts. For (**A**), schematic depicts blood sampling across gestation and plasma isolation. X-axis represents gestational age during pregnancy and time post-delivery (weeks) thereafter (ns = not significant, * p < 0.05 ** ≤ 10^−7^). BMI data is not available for Validation 2.

For all cohorts, we included individuals of diverse racial and ethnic backgrounds in approximately matched proportions across NT and PE groups (Table S1). We recorded maternal pre-pregnancy and pregnancy characteristics, and defined a pregnancy as NT if it was both uncomplicated and went to full-term (≥ 37 weeks) or as PE with or without severe features based on current guidelines (see Methods). For mothers who developed PE, all antenatal blood samples were collected prior to diagnosis.

Our final analysis included a subset of those samples which passed pre-defined quality metrics (Supp. note 1, Methods, Fig S1). After confirming sample quality, 404 samples from 199 mothers (142 NT, 57 with PE) were included in the final analysis (Table S2). Specifically, 209, 106, and 89 samples from 73, 39, and 87 participants (49, 32, 61 NT; 24, 7, 26 with PE) were included in Discovery, Validation 1, and Validation 2, respectively (RNAseq).

Across gestational timepoints in all cohorts, we found no significant difference in sampling time between PE and NT groups (p ≥ 0.26, 0.11, 0.46). Known risk factors for PE, such as pre-pregnancy maternal body mass index (BMI), maternal age, and gravidity followed expected trends. BMI specifically was significantly different between PE and NT groups in Discovery cohort alone (p = 0.02, 0.45, NA) while maternal age and gravidity were not (p ≥ 0.29, 0.16, 0.2) (Fig 1B, Table S1). History of PTB and mode of delivery were significantly different between NT and PE groups only for Validation 2. Other demographic factors like race, ethnicity, and nulliparity differed across cohorts but not between case groups within each cohort (adjusted p ≤ 0.05, chi-squared test for categorical or ANOVA for continuous variables with Bonferroni correction, Table S1).

In mothers who later developed PE, we observed no significant difference (p = 0.14, 1.0, 0.4) in gestational age at onset between those who did not experience severe symptoms (n = 11, 4, 3*) as compared with those with did experience severe (n = 13, 3, 13*) symptoms (Fig 1C). Furthermore, 21 mothers who developed PE also delivered preterm (n = 9, 1, 11) as compared with no mothers in the NT group as reflected by significantly different gestational ages at delivery (p = 10^−7^, 0.04, 10^−9^, one-sided test) (Fig 1D) and lower fetal weight at delivery (Table S1), which was consistent with epidemiological evidence that PE increases the risk of spontaneous or indicated preterm delivery ^5,27^ (values reported as Discovery, Validation 1, Validation 2, Mann-Whitney rank test unless otherwise specified, * denotes data were incomplete for specified cohort).

### Identification of cfRNA changes across gestation in mothers who developed PE

There were 544 differentially expressed genes (DEGs) that differed across gestation and post-partum between mothers who later developed PE with or without severe features as compared with NT mothers who did not experience complications (adjusted p ≤ 0.05, see Methods and Supplementary Note 2). Most DEGs (498) were annotated as protein-coding, and a small fraction (43, 8%) were other types including 11 mitochondrial t-RNAs, 6 long non-coding RNAs, 8 pseudogenes, and 1 small nucleolar RNA (snoRNA).

To further understand when these changes occur during gestation, we estimated the log fold-change (logFC) for each gene by each gestational time point as well as post-partum. We observed that these gene changes occurred most strikingly before 20 weeks of gestation as indicated by a clear bimodal distribution with two peaks centered around logFC of +0.8 and -0.6 (Fig 2A). We also found that gene changes were also most stable before 20 weeks of gestation where over 50% of genes had a coefficient of variation (CV) < 1 as compared to 31% at or after 23 weeks of gestation and 36% at post-partum (Fig S2A).

**Figure 2.**
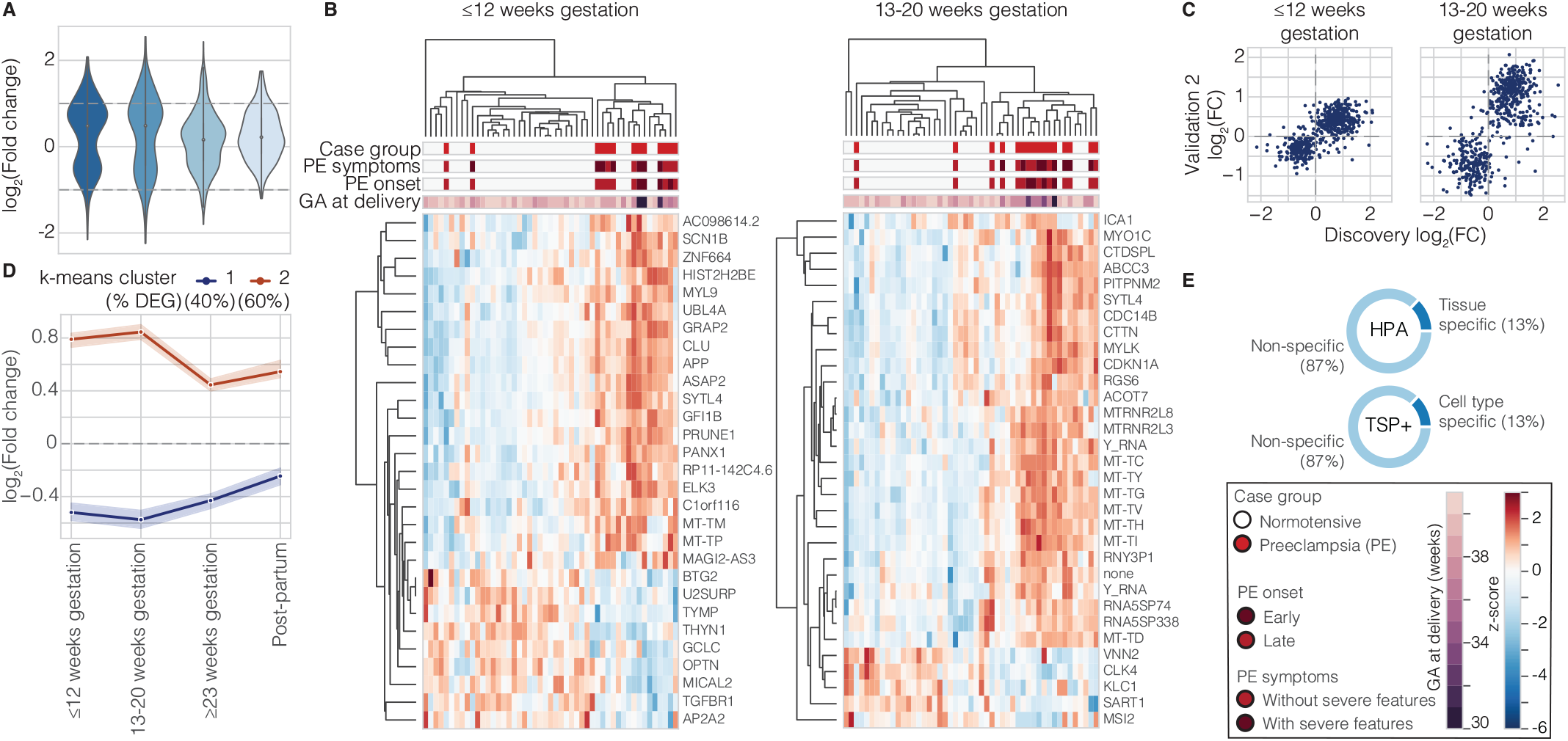
Before 20 weeks of gestation, changes in the cfRNA transcriptome segregate PE and NT samples and are enriched for neuromuscular, endothelial and immune cell types and tissues. (**A**) Distribution of log(Fold change) with dashed lines at log(Fold change) = ± 1 (**B**) At ≤12 and between 13–20 weeks of gestation, a subset of differentially expressed genes can separate PE and NT samples despite differences in symptom severity, PE onset subtype, and gestational age (GA) at delivery. (**C**) Comparison of log(Fold change) for DEGs between Discovery (x-axis) and Validation 2 (y-axis) reveals excellent agreement: 92%, and 94% of genes had the same logFC sign with a spearman correlation of 0.71 and 0.72 (p < 10^−15^) at ≤12 and 13–20 weeks of gestation, respectively. (**D)** Across gestation, differentially expressed genes for PE as compared to NT can be described by 2 longitudinal trends: Increased in PE over gestation in orange and Decreased in PE over gestation in dark blue as revealed by k-means clustering. (**E**) Approximately 13% of DEGs are tissue- or cell-type specific when compared with the Human Protein Atlas (HPA) and an augmented Tabula Sapiens (TSP+) atlas.

We then asked whether a subset of gene changes approximately proportional in number to total sample number (n = 49, 49, 57, 46 for each time point) was sufficient to segregate PE (n = 13, 16, 20, 17) and NT (n = 36, 33, 37, 29) samples across gestation. We found that 24-32 genes were sufficient to separated PE and NT samples across gestation and at post-partum with good specificity (86% [75–93%], 79% [66–88%], 97% [90–100%], and 90% [78–96%]) and sensitivity (85% [64–95%], 88% [69–96%], 65% [47–80%], and 71% [51–86%]) (Fig 2B, S2B, All values reported for ≤12 weeks, 13-20 weeks, ≥23 weeks of gestation and post-partum, [90% CI]) (See also Fig S3, File S1).

Nearly all 544 DEG changes showed excellent agreement in both Validation cohorts as compared to Discovery across gestation but not post-partum. Specifically, more than 82% and 92% of genes across gestation had the same logFC sign with a spearman correlation of at least 0.67 and 0.71 for Validation 1 and 2 respectively (p<10^−15^) as compared with 60% and 0.35 post-partum (Fig 2C, S2C). Finally, we asked whether symptom severity (without or with severe features) correlated with logFC magnitude for these 544 DEGs common to both PE subtypes. We found that on average, symptom severity did not influence logFC magnitude as reflected by a slope of nearly 1 across gestation and post-partum (Fig S2E).

### cfRNA changes reflect disease pathophysiology in mothers who developed PE

The 544 identified DEGs could be well categorized into two longitudinal trends (Fig 2D, S4A,C). Resembling a valley or V-shape, the first trend (Group 1) described the longitudinal behavior of 216 genes (40%), for which measured levels were reduced in PE samples (−1.3x to -1.5x) across gestation with a minimum between 13–20 weeks. Peaking in early gestation before 20 weeks (1.75x), the second trend (Group 2) described the behavior of 328 genes (60%) that had significantly elevated levels in PE samples before 20 weeks and to a lesser extent after 23 weeks of gestation (1.3x). For Group 1 but not 2, gene changes were far less evident post-partum and trended toward no difference between PE and NT, which may reflect a placental contribution to DEG levels.

Approximately 13% of DEGs were tissue or cell-type specific (Fig 2E). Genes that were decreased in PE across gestation (Group 1) were broadly enriched for the immune system, whereas, those genes increased in PE across gestation (Group 2) were enriched for nervous, muscular, endothelial, and immune contributions as reflected by cell-type (adjusted p ≤ 0.05, hypergeometric test) and pathway enrichment (adjusted p ≤ 0.05, hypergeometric test) (Fig S2E, Table S3). Consistent with current understanding of the pathogenesis of PE, we identified a strong endothelial-linked signal underscored by contributions from capillary aerocytes (p = 0.03), an endothelial cell-type specific to the lungs ^45^, platelets (p = 10^−33^), and several platelet related pathways like platelet degranulation and platelet activation, signaling, and aggregation (p ≤ 10^−8^) among others.

Surprisingly, we also found significant, elevated nervous and muscular contributions for PE as emphasized by contributions from excitatory neurons (p = 0.02), oligodendrocytes (p = 0.005), and smooth muscle (p = 0.0003) and terms like muscle contraction and dilated cardiomyopathy (p ≤ 0.02). The immune system also contributes to both elevated (e.g., mesenchymal stem cells, total PBMCs) and decreased (granulocytes, T-cells) changes across gestation. Genes in both groups were enriched for signaling pathways (i.e., secretion by cell, integrin-mediated signaling pathway, regulation of I-kappaB kinase, NF-kappaB signaling). Group 2 was also enriched for cellular compartments such as the cell periphery, cell junctions, and extracellular space, consistent with reports that PE may be associated with signaling from the fetoplacental complex ^46^. Finally, DEGs were broached enriched for genes previously implicated in PE ^47^ (30 gene overlap, p = 0.006, hypergeometric test) (Table S3).

### A machine-learning classifier predicts risk of PE on or before 16 weeks of gestation

Since gene changes associated with PE pathogenesis across gestation are readily detected irrespective of symptom severity, we sought to build a classifier that could identify mothers at risk of PE at or before 16 weeks of gestation (Supplementary Note 2, Fig S2E). We trained a logistic regression model on the Discovery cohort (n = 61 NT, 24 PE samples). After training, the final model performed well with a near perfect AUROC (0.99 [0.99–0.99]), good specificity (85% [77– 91%]), and perfect sensitivity (100% [92-100%]) (Fig 3A, Table 1). We then tested this model on Validation 1 (n = 35 NT, 8 PE) and two other independent cohorts, which were collected at separate institutions: Validation 2 (n = 61 NT, 28 PE samples) and Del Vecchio and colleagues ^33^ cohort (n = 8 NT, 5 with PE, 7 with gestational diabetes, 2 with chronic hypertension). Across these cohorts, the final model once again performed well with consistent AUROC (0.71 [0.70-0.71], 0.72 [0.71– 0.72], 0.74 [0.73–0.74]), sensitivity (75% [46–92%], 56% [42-72%], 60% [26–87%]) and specificity (56% [43–70%], 69% [59-78%], 100% [89–100%]) (All reported as Validation 1, Validation 2, Del Vecchio) (Fig 3A, Table 1).

**Table 1.** PE prediction performance metrics for samples collected early in gestation (between 5–16 weeks). Control and case sample numbers are reported as the total sample number and in parentheses, the number of samples misclassified. All other statistics including sensitivity specificity, PPV, NPV, and AUROC are reported as the estimated percentage followed by the 90% CI in square brackets. In Del Vecchio^1^, the control group is defined as samples from any pregnant mother who did not develop PE including those with other underlying or pregnancy-related complications like chronic hypertension and gestational diabetes respectively. In Del Vecchio^2^, the control group is defined as samples strictly from NT pregnant mothers who did not experience complications.

|  | **Control**<br>**(n)** | **Case**<br>**(n)** | **Sensitivity**<br>**(%)** | **Specificity**<br>**(%)** | **PPV**<br>**(%)** | **NPV**<br>**(%)** | **AUC** |
| --- | --- | --- | --- | --- | --- | --- | --- |
| Discovery | 61 (9) | 24 (0) | 100<br>[92-100] | 85<br>[77-91] | 73<br>[59-84] | 100<br>[96-100] | 0.99<br>[0.99-0.99] |
| Validation<br>1 | 35 (11) | 8 (3) | 75<br>[46-92] | 56<br>[43-70] | 28<br>[15-46] | 91<br>[77-97] | 0.71<br>[0.7-0.71] |
| Validation<br>2 | 61 (19) | 28 (12) | 56<br>[42-71] | 69<br>[59-78] | 46<br>[32-59] | 78<br>[68-86] | 0.72<br>[0.71-0.72] |
| Del<br>Vecchio<sup>1</sup><br>(2020) | 17 (0) | 5 (2) | 60<br>[26-87] | 100<br>[89-100] | 100<br>[56-100] | 89<br>[74-97] | 0.74<br>[0.73-0.74] |
| Del<br>Vecchio<sup>2</sup><br>(2020) | 8 (0) | 5 (2) | 60<br>[26-87] | 100<br>[79-100] | 100<br>[56-100] | 80<br>[55-94] | 0.80<br>[0.79-0.80] |

**Figure 3.**
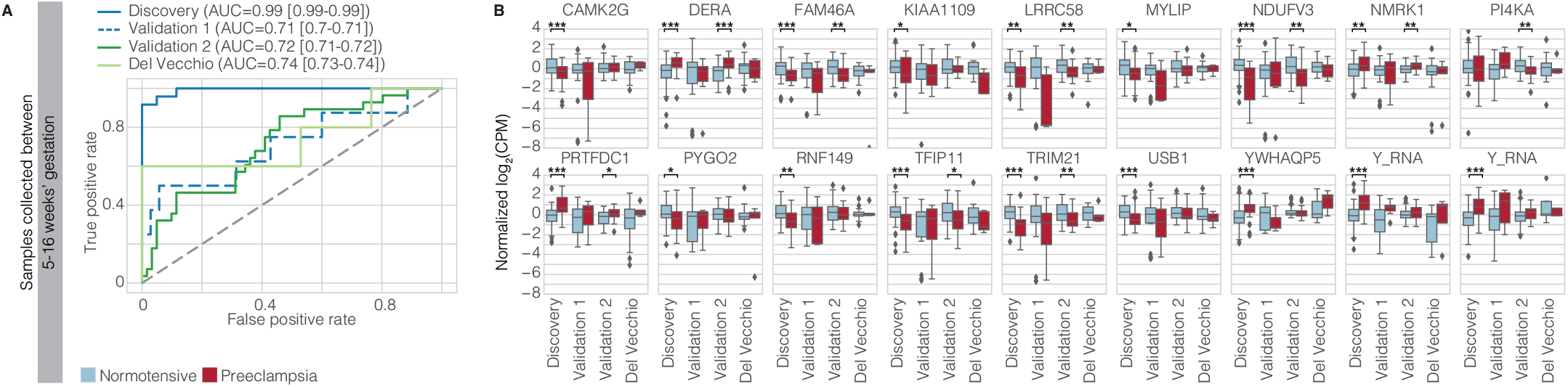
A subset of cfRNA changes can predict risk of PE early in gestation. (**A**) Classifier performance as quantified by ROC for samples collected in early gestation between 5–16 weeks. For each cohort, including 3 validation cohorts of which Validation 2 and Del Vecchio are independent, the legend states the AUROC and the corresponding 90% CI in square brackets. (**B**) Prediction of PE incorporates cfRNA levels for 18 genes for which normalized centered log2(Fold change) trends hold across discovery, Validation 1, Validation 2, and Del Vecchio cohorts as confirmed using univariate analysis (*p ≤ 0.05, **p ≤ 0.01, ***p ≤ 0.005; one-sided Mann-Whitney U-Test with Benjamini-Hochberg correction). For box-plots, center line, box limits, whiskers, and outliers represent the median, upper and lower quartiles, 1.5x interquartile range, and any outliers outside that distribution respectively. Plot limits are -8 to 4 to better visualize the main distribution.

Next, we inspected erroneously classified samples from the Validation 2 and Del Vecchio cohorts. For false negatives in Validation 2 and Del Vecchio, we find a shift to later gestational ages at collection (13.5 ± 2, 12.5 ± 2 weeks) as compared to PE samples that were correctly classified (12 ± 2, 12 ± 0 weeks; mean ± SD for Validation 2, Del Vecchio) (Fig S5A). This suggests that in practice, there may an optimal collection window to reduce false negatives. Indeed, if we only consider samples before 14 weeks of gestation, we observe a 9% and 15% increase in sensitivity with corresponding AUROC values of 0.73 and 0.90 for Validation 2 and Del Vecchio respectively. There were no false positives in the Del Vecchio cohort, suggesting that the model can distinguish between PE and other risks like chronic hypertension or gestational diabetes. The model also proved well-calibrated estimating a slightly elevated probability of PE for gestational diabetes (0.15 ± 0.08) and chronic hypertension (0.18 ± 0.13) – known PE risk factors ^48–54^ – as compared to the estimate for NT samples (0.09 ± 0.08) (mean ± SD, Fig S5B). These elevated probabilities for other risk factors impacted the test’s AUROC (0.74 [0.73-0.74] as compared to 0.8 [0.79-0.8] for only PE vs NT samples, Table 1) (all reported as value, [90% CI]).

Finally, we inspected the 18 genes (Fig 3B, Table S4) used by the model to yield probability estimates. Eight genes were annotated in the Human Protein Atlas (HPA)^55^ as enhanced or enriched in the placenta (FAM46A, MYLIP), neuromuscular (CAMK2G, NDUFV3, PI4KA, PRTFDC1) and immune systems (RNF149, TRIM21). Univariate analysis further confirmed that 9 of the gene trends (i.e., decreased or increased gene levels in PE) observed in the Discovery dataset are upheld in Validation 2 (adjusted p ≤ 0.05, one-sided Mann-Whitney rank test) (Fig 3B). We also found that the majority of models trained using a subset of the 18 initial genes can predict future PE onset with varying performance. Notably, performance improved across all metrics (sensitivity, specificity, and AUROC) as we increased the number of genes included for model training (Table S5, Fig S5C).

### The multifactorial nature of PE and maternal organ health is reflected in cfRNA

We next wondered whether cfRNA measurement reflects the multifactorial nature of PE. We identified 503 DEGs (adjusted p ≤ 0.05) that differed across gestation between mothers who later developed PE with as compared to without severe symptoms. Since there were no significant differences in symptom severity as related to timing of PE onset (Fig 1C), we believe that our observations contrasting PE with and without severe symptoms are not obscured by differences in PE-onset type. As before, most DEGs (484) were annotated as protein-coding, and a small fraction (18, 4%) were other types including 12 pseudogenes and 4 long non-coding RNAs.

We observed that DEGs could be well categorized into 4 longitudinal trends (Fig 4A, S4B,D). Two groups (Group 1, 3) described the temporal behavior of 217 genes (44%), for which measured levels were either consistently increased (Group 1) or reduced (Group 3) in PE with as compared to without severe symptoms (±1.8x) across gestation and trended towards no change post-partum. In contrast, Groups 2 and 4 (286 genes, 56%) changed signs in mid-gestation beginning as slightly elevated (Group 2, 1.2x) or decreased (Group 4, -1.2x) in severe PE and then moving to decreased (Group 2, -1.4x) or unchanged (Group 4, 1x) at ≥23 weeks of gestation.

**Figure 4.**
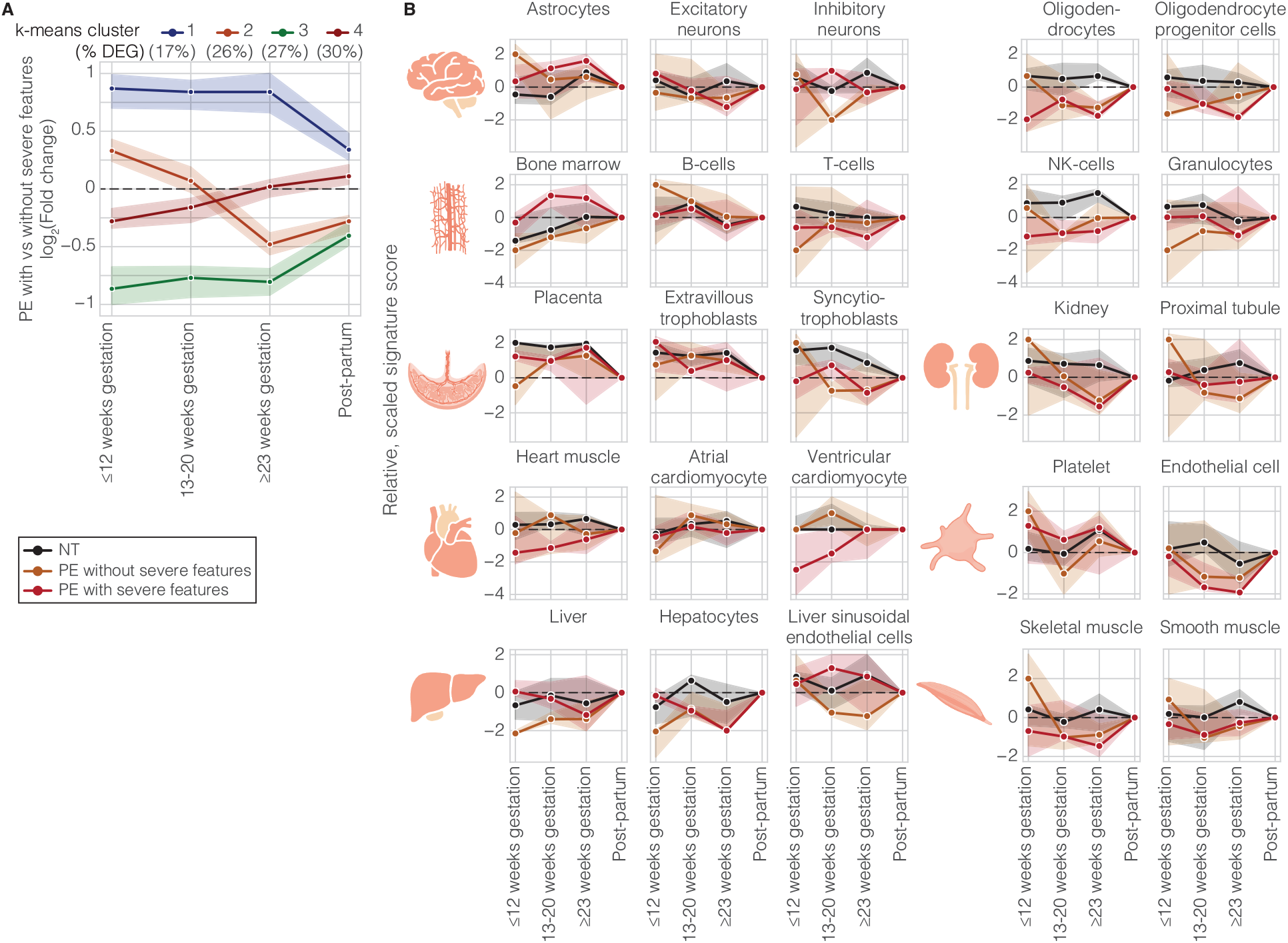
Changes in the cfRNA transcriptome reflect PE’s multifactorial nature and pathogenesis over pregnancy prior to diagnosis. (**A**) Across gestation, differentially expressed genes for PE with as compared to without severe features (503 DEGs) can be described by 4 longitudinal trends as revealed by k-means clustering. Points indicate median per DEG cluster and shaded region indicates 95% CI. (**B**) Comparison of organ and cell-type changes over gestation for eight organ systems reflect the multifactorial nature of PE and provide a possible means to monitor maternal organ health (Top to bottom: Brain, immune, placenta and kidney, heart and endothelial-linked, liver and muscle). Points indicate median per sample group (NT in black, PE without severe features in yellow, PE with severe features in red) and shaded region indicates 75% CI.

Analysis of the enriched cell types and tissues of origin for each of these groups revealed that elevated gene differences in severe PE were driven by contributions from endothelial cells and the adaptive immune system (bone marrow). In contrast, genes that changed signs over gestation were enriched for innate immune cell types (e.g., granulocytes and neutrophils for Group 2, thymus for Group 4) (Table S6). Quantifying total cfRNA signal confirmed an increased bone marrow signal for only severe PE across gestation and a decreased granulocyte signal for only PE without severe features at ≤12 weeks of gestation (Fig 4B). Gene ontology analysis further revealed pathways specific to genes that were only decreased for severe PE in early gestation (Group 4) like axon guidance, nervous development, and metabolism of RNA (adjusted p < 0.05).

We next investigated whether it might be possible to monitor organ health noninvasively focusing on eight organ systems (Fig 4B) relevant to PE presentation with consequences such as proteinuria, impaired liver function, renal insufficiency, and epilepsy. We found striking shifts in total contributions for all systems. We observed an increased astrocyte signal before 20 weeks of gestation and decreased oligodendrocytes and excitatory neurons at ≥23 weeks of gestation for all PE relative to NT (Fig 4B, 1^st^ row). Although placental contributions increased over pregnancy with a peak in late gestation as expected, placental tissue and syncytiotrophoblast contributions were reduced for PE pregnancies before 20 weeks of gestation. Finally, we observed a decreased signal in hepatocyte, kidney, endothelial cell, and smooth muscle signatures across gestation and increased platelet signal before 12 weeks of gestation for PE. These tissue and cell type specific changes are both consistent with common PE pathogenesis and the specific, prominent diagnoses in our cohort (e.g., thrombocytopenia, proteinuria, impaired liver function, renal insufficiency).

## Discussion

Noninvasive measurements of the cf-transcriptome present an opportunity to study human development and disease from any organ at a molecular scale. Here, we showed that circulating cfRNA measurements taken during pregnancy can clearly distinguish between PE and NT pregnancies. We found that the most striking differences occur early on and broadly showed no reproducible difference at post-partum, consistent with known PE etiology, namely that the placenta drives the disease process ^34,36^. We next validated these changes using two separate cohorts (Validation 1 and 2) and explored their physiological relevance.

Our findings provide molecular evidence supporting generally accepted physiological understanding of PE pathogenesis: early abnormal placentation and systemic endothelial dysfunction ^34^. Early in gestation, we observe a reduced placental signal for PE regardless of onset type or symptom severity. Concurrently, platelets and endothelial cells drive cfRNA changes in all PE as compared to NT and between PE with or without severe symptoms especially before 20 weeks of gestation. Increases in cell-type specific cfRNA may occur through signaling and secretion by cells, as underscored by functional enrichment analysis. The innate and adaptive immune system also heavily contribute to cfRNA changes in PE with clear, marked shifts related to bone marrow, T-cells, B-cells, granulocytes, and neutrophils, consistent with previous studies on the maternal-placental interface and PE ^34,56–58^.

PE is a broad and complex syndrome. Because the complication can present clinically across more than 20 weeks with a diversity of symptoms, significant effort has been made to subclassify the disease based on timing of onset. Used as a proxy for hypothesized pathogenesis, the timing of PE onset subdivides the disease into ‘placental’ early-onset PE (occurring before 34 weeks of gestation) and ‘maternal’ late-onset PE (occurring on or after 34 weeks) ^34,37,59^. However, early- and late-onset PE may represent a spectrum of disease severity that corresponds with timing of onset and may lead to additional pregnancy complications, such as intrauterine growth restriction (IUGR) ^34,36,38,39^. Indeed, some evidence such as transcriptional profiling of whole blood drawn at diagnosis suggests a common gene signature for both subtypes ^40^. Our findings further corroborate a common signature across onset types and suggest that PE may be better stratified based on symptom severity.

Indeed, PE may be best subtyped molecularly. Given the diversity of clinical presentations, we propose a novel, noninvasive means of monitoring a mother’s risk of specific organ damage, common in PE. The cfRNA changes we characterized here reflect dysfunction in at least five organ systems (brain, liver, kidney, muscle, bone marrow), and can in some cases further distinguish between PE with or without severe symptoms. As a molecular lens into maternal health, liquid biopsies present an opportunity as both a research and clinical tool to learn about the pathogenesis of a human disease in humans and as a predictor of maternal health. Here, we have shown proof of principle that cfRNA measurements can form the basis for a robust liquid biopsy test, which predicts PE very early in gestation and if validated in controlled clinical studies, could help discover and manage those at risk for PE. Such a test could serve as a complement to recent efforts based on clinical and laboratory data^60^, and even be coupled with tests taken later during gestation. We further demonstrated that cfRNA measurements reflect who is at risk for specific organ damage. Together, these results form the basis for a series of clinical tests that can be used to help characterize and stratify the pathogenesis of PE in real time, to date unrealized key objectives for obstetric care.

## Data Availability

Raw and processed sequencing data will be deposited with the SRA and GEO, respectively.

## Acknowledgements

We thank N. Aghaeepour and I. Maric from Stanford University for helpful discussion and data analysis suggestions, the many researchers and clinicians affiliated with the March of Dimes Prematurity Research Center at Stanford University for insightful feedback after oral presentations, and the participants in this study for their invaluable contribution.

## Author contributions

M.N.M., G.M.S., D.K.S., and S.R.Q. conceptualized and designed this study. M.L.D., V.D.W., G.M.S., and D.K.S. designed the cohort study at Lucile Packard Children’s Hospital. R.J.W., A.A.C., and C.C.Q. collected and processed whole blood samples and corresponding sample and participant metadata. GAPPS oversaw, collected, and processed whole blood samples and corresponding sample and participant metadata at GAPPS. M.N.M. developed experimental protocols for cfRNA extraction and data generation and processed all samples from cfRNA extraction up to and including RT-qPCR and library generation for RNA sequencing. R.V.S., M.T., A.D., H.M., and N.F.N. developed and executed experimental protocols for RNA sequencing and computational protocols for raw sequencing data transfer. S.K.V. developed computational methods to define genes as cell-type and tissue specific in the context of the whole body. M.N.M and S.K.V. designed the analyses to characterize tissue and cell type contributions. M.N.M. conceptualized and developed computational analyses, analyzed the data, and wrote the initial manuscript draft in collaboration with S.R.Q.. All authors contributed to the writing and editing the manuscript.

## Materials & methods

### Clinical study design

Discovery and Validation 1 were collected as part of a longitudinal, prospective study. We enrolled pregnant mothers (aged 18 years or older) receiving routine antenatal care on or prior to 12 weeks of gestation at Lucile Packard Children’s Hospital at Stanford University, following study review and approval by the Institutional Review Board (IRB) at Stanford University (#21956). All signed informed consent prior to enrollment. Whole blood samples for plasma isolation were then collected at three distinct timepoints during their pregnancy course and once (or twice for 2 individuals) post-partum.

Validation 2 was collected as part of the Global Alliance to Prevent Preterm and Stillbirth (GAPPS) at several, independent sites. Samples were processed and sequenced at Stanford under the same IRB as above (#21956). All signed informed consent prior to enrollment. Whole blood samples for plasma isolation were collected at a single time point (or 2 timepoints in the case of 2 individuals with PE) prior to or at 16 weeks of gestation.

For all three cohorts, we ensured that all included individuals did not have chronic hypertension or gestational diabetes. Mothers were defined as having PE based upon current American College of Obstetrics and Gynecology (ACOG) guidelines (see below). Mothers were defined as controls if they had uncomplicated term pregnancies and either normal spontaneous vaginal or caesarean deliveries. For mothers who developed PE, all antenatal samples included in this study were collected prior to clinical diagnosis.

We tested for within cohort (NT vs PE) and across cohort differences in demographic variables using a chi-squared test and ANOVA for categorical and continuous variables respectively. We then applied Bonferroni correction and reported any differences as significant if adjusted p ≤ 0.05.

### Definition of PE

PE was defined per the ACOG guidelines ^27^ based on two diagnostic criteria: 1) new-onset hypertension developing on or after 20 weeks of gestation and 2) new-onset proteinuria or in its absence, thrombocytopenia, impaired liver function, renal insufficiency, pulmonary edema, or cerebral or visual disturbances.

New-onset hypertension was defined when the systolic and/or diastolic blood were at least 140 or 90 mmHg, respectively, on at least 2 separate occasions between 4 hours and 1 week apart. Proteinuria was defined when either 300 mg protein was present within a 24-hour urine collection or an individual urine sample contained a protein/creatinine ratio of 0.3 mg/dL, or if these were not available, a random urine specimen had more than 1 mg protein as measured by dipstick. Thrombocytopenia, impaired liver function, and renal insufficiency were defined as a platelet count of < 100,000/µL, liver transaminases ≥ 2x of normal, and serum creatinine > 1.1 mg/dL, respectively.

Symptoms were defined as severe per the ACOG guidelines. Specifically, PE is defined as severe if any of the following symptoms were present and diagnosed as described above: new-onset hypertension with systolic and/or diastolic blood pressure of at least 160 or 110 mmHg respectively, thrombocytopenia, impaired liver function, renal insufficiency, pulmonary edema, new-onset headache unresponsive to medication and unaccounted for otherwise, or visual disturbances.

Finally, a pregnant mother was considered to have early-onset PE if onset occurred before 34 weeks of gestation and late onset thereafter.

### Sample preparation

#### Plasma processing

At Lucile Packard Children’s Hospital, blood samples were collected in either EDTA-coated (Cat No 368661, Becton-Dickinson) or Streck cfRNA BCT (Cat No 218976, Streck) tubes at ≤12, 13– 20, and ≥23 weeks of gestation, and post-partum for each participant. Within 30 minutes, tubes were then centrifuged at 1600g for 30 minutes at room temperature. Plasma was transferred to 2-mL microfuge tubes and centrifuged at 13000g for another 10 minutes in a microfuge. One milliliter aliquots were then transferred to 2-mL Sarstedt screw cap microtubes (Cat No 50809242, Fisher Scientific) and stored at -80°C until analysis.

At GAPPS, blood samples were collected in EDTA-coated tubes at ≤16 weeks of gestation from a network of collection sites including Yakima Valley Memorial Hospital, Swedish Medical Center, and the University of Washington Medical Center. Per Standard Operating Procedure (SOP), tubes were then centrifuged within 2 hours of collection at 2500RPM for 10 minutes at room temperature in a swinging bucket rotor. Plasma was transferred to 2-mL cryovials in at most 1 mL aliquots and stored at -80°C until analysis. Sample volume was also recorded.

#### cfRNA isolation

In 96-sample batches, cfRNA from 1-mL plasma samples was extracted in a semi-automated fashion using the Opentrons 1.0 system and Norgen Plasma/Serum Circulating and Exosomal RNA Purification 96-Well Kit (Slurry Format) (Cat No 29500, Norgen). Samples were subsequently treated with Baseline-ZERO DNAse (Cat No DB0715K, Lucigen) for 20 minutes at 37°C. DNAse-treated cfRNA was then cleaned and concentrated into 12 µL using Zymo RNA Clean and Concentrator-96 kits (Cat No R1080).

Following cfRNA extraction from plasma samples, isolated RNA concentrations were estimated for a randomly selected 11 samples per batch using Bioanalyzer RNA 6000 Pico Kit (Cat No 5067-1513, Agilent) per manufacturer instructions.

#### Sequencing library preparation

cfRNA sequencing libraries were prepared with SMARTer Stranded Total RNAseq Kit v2 - Pico Input Mammalian Components (Cat No 634419, Takara) from 4 µL of eluted cfRNA according to the manufacturer’s instructions. Samples were barcoded using SMARTer RNA Unique Dual Index Kit – 96U Set A (Cat No 634452, Takara), and then pooled in an equimolar fashion and sequenced on Illumina’s NovaSeq platform (2×75 bp) to a mean depth of 54, 33, and 38 million reads per sample for Discovery, Validation 1, and Validation 2 cohorts, respectively. Some samples (12, 61, 0 for Discovery, Validation 1, and Validation 2 cohorts) were not sequenced due to failed library preparation.

### Sequencing data analysis

#### Bioinformatic processing

For each sample, raw sequencing reads were trimmed using Trimmomatic (v 0.36) and then mapped to the human reference genome (hg38) with STAR (v 2.7.3a). Duplicate reads were then removed by GATK’s (v 4.1.1) MarkDuplicates tool. Finally, mapped reads were sorted and quantified using htseq-count (v 0.11.1) generating a counts table (genes x samples). Read statistics were estimated using FastQC (v 0.11.8).

Across samples, the bioinformatic pipeline was managed using Snakemake (v 5.8.1). Read and tool performance statistics were aggregated across samples and steps using MultiQC (v 1.7). Following sample quality and gene filtering, all gene counts were adjusted to log2-transformed counts per million reads (CPM) with trimmed mean of M values (TMM) normalization ^61^.

#### Sample quality filtering

For every sequenced sample, we estimated three quality parameters as previously described ^62,63^. To estimate RNA degradation in each sample, we first counted the number of reads per exon and then annotated each exon with its corresponding gene ID and exon number using htseq-count. Using these annotations, we measured the frequency of genes for which all reads mapped exclusively to the 3’ most exon as compared to the total number of genes detected. RNA degradation for a given sample can then be approximated as the fraction of genes where all reads mapped to the 3’ most exon. To estimate the number of reads that mapped to genes, we summed counts for all genes per sample using the counts table generated from bioinformatic processing above. To estimate DNA contamination, we quantified the ratio of reads that mapped to intronic as compared to exonic regions of the genome.

After measuring these three metrics across nearly 700 samples, we empirically estimated RNA degradation and DNA contamination’s 95^th^ percentile bound. We considered any given sample an outlier, low quality sample if its value for at least one of these metrics was greater than or equal to the 95^th^ percentile bound or if no reads were assigned to genes.

Once values for each metric were estimated across the entire dataset, we visualized: (1) whether low-quality samples clustered separately using hierarchical clustering (average linkage, Euclidean distance metric) and (2) whether sample quality drove variance in gene measurements using principal component analysis (PCA). These analyses were performed in Python (v 3.6) using Scikit-learn for PCA (v 0.23.2), Scipy for hierarchical clustering (v 1.5.1), and nheatmap for heatmap and clustering visualization (v 0.1.4).

#### Gene filtering

We performed filtering to identify well-detected genes across the entire cohort. Specifically, we used a basic cutoff that required a given gene be detected at a level of at least 0.5 CPM in at least 75% of discovery samples after removing outlier samples. Following this step, we retain 7,160 genes for DE analysis.

#### Differential expression analysis

Differential expression analysis was performed in R using Limma (v 3.38.3). To identify gene changes associated with PE across gestation and post-partum, we used a mixed-effects model. We performed DE using two design matrices: (1) Examine the interaction between time to PE onset or delivery for NT and PE symptoms (i.e., PE with or without severe symptoms) and (2) Examine the interaction between time to PE onset or delivery for NT and PE broadly. In both design matrices, we included time to PE onset or delivery for NT (continuous variable), whether a sample was collected post-partum (binary variable), the interaction between time and PE symptoms for (1) or PE for (2), the interaction between whether a sample is post-partum and PE symptoms for (1) and PE for (2), and 7-8 confounding factors.

In (1), we defined PE symptoms categorially using 3 levels - NT, PE without severe symptoms, PE with severe symptoms). In (2), we defined whether a sample was PE using a binary, indicator variable (0 = NT, 1 = PE). The 7-8 confounding variables included were maternal race (categorial variable), maternal ethnicity (binary variable), fetal sex (binary variable), maternal pre-pregnancy BMI group (categorical variable), maternal age (continuous variable, only included in design 1), and sequencing batch (categorical variable). We defined time to PE onset or delivery as the difference between gestational age at onset or delivery and gestational age at sample collection. We defined BMI group as follows: Obese (BMI ≥ 30), Overweight (25 ≤ BMI < 30), Healthy (18.5 ≤ BMI < 25), Underweight (BMI < 18.5). We chose to model time to PE onset or delivery as a continuous variable, specifically a natural cubic spline with 4 degrees of freedom to account for the range across which samples were collected (1–3 months per collection period). We also blocked for participant identity (categorical variable), modeling it as a random effect to account for auto-correlation between samples from the same person.

Per the Limma-Voom guide, to account for sample auto-correlation over time, we ran the function voomWithQualityWeights twice. We first ran it without any blocking on participant identity, and used this base estimation to approximate sample auto-correlation based on participant identity using the function duplicateCorrelation. After estimating correlation, voomWithQualityWeights was run again, this time blocking for participant identity and including the estimated auto-correlation level. A linear model was then fit for each gene using lmFit and differential expression statistics were approximated using Empirical Bayes (eBayes) methods. For comparing PE with vs without severe symptoms, we contrasted the relevant coefficients (makeContrasts) and then applied Empirical Bayes as opposed to directly after lmFit.

DEGs were then identified using the relevant design matrix coefficients and the function, topTable, with Benjamini-Hochberg multiple hypothesis correction at a significance level of 0.05. For design 1, we identified DEGs related to 3 comparisons: PE without severe symptoms vs NT (1759 DEGs), severe PE vs NT (1198 DEGs), and PE with vs without severe symptoms (503 DEGs). We find 544 genes in common for PE without and with severe symptoms vs NT. These 544 DEGs were explored in Figure 2 and the related main text. For design 2, we identified DEGs related to PE vs NT alone (330 DEGs), which we used as the initial gene set for building a logistic regression model (see Supplementary Note 2). Finally, we removed the effect of sequencing batch alone on estimated logCPM values with TMM normalization for the Discovery cohort using the limma-voom function, removeBatchEffect.

#### log(Fold change) and CV estimation

We define log2-transformed fold-change (logFC) as the difference between the median gene level (logCPM, see Bioinformatic processing section above) between PE and NT samples for a given sample collection period (i.e., ≤12, 13–20, and ≥23 weeks of gestation, or post-partum). In the case where a given person had multiple samples included into a specific collection period, we only used the values associated with the first collected sample to avoid artificially reducing within-group (PE or NT) variance due to auto-correlation among samples from the same person.

We then quantified the relative dispersion around the estimated logFC for each gene using an approximation for CV. Specifically, we consider CV to be the ratio between an error bound, ∂, and the estimated logFC. We defined the error bound, ∂, as the one-sided error bound associated with the lower (or upper in the case of negative logFC values) 95% CI as estimated by bootstrapping. This definition of ∂ as a one-sided bound that approaches 0 (equivalent to no FC) allowed us to explore how confident we could be in an estimated logFC. For instance, a CV = 1 would indicate that at the boundary of proposed values, the logFC for a given gene becomes effectively 0. Similarly, a CV > 1 would indicate even less confidence in a proposed average logFC and indicate that at the boundary, the estimated logFC changes signs (i.e., a negative logFC becomes a positive one or vice versa).

#### Hierarchical clustering analysis

For each sample collection period, hierarchical clustering was performed using differentially expressed genes with an |logFC| ≥ 1 and CV < 0.5 or 0.4 in the case of the 13–20 weeks of gestation time point so that the number of genes used did not exceed the number of samples. For each gene that passed these thresholds, we calculated a z-score across all samples (at most 1 per subject, the earliest collected sample in a given group) in each sample collection period using the function StandardScaler in Scikit-learn (v 0.23.2), Average linkage hierarchical clustering with a Euclidean distance metric was then performed for both rows (gene z-scores) and columns (samples in same collection group) for a given matrix in Python using Scipy (v 1.5.1). Clustering and corresponding heatmaps were visualized using nheatmap (v 0.1.4).

#### Longitudinal dynamics analysis

To group gene changes by longitudinal behavior, we performed k-means clustering on a matrix where each row was a differentially expressed gene and each column was the estimated logFC for a given sample collection period (N genes x 4 timepoints). We measured the sum of squared distances for every ‘k’ between 1 and 16 (4^2^) where 16 represents the maximum possible k (4 timepoints with 2 possibilities each, logFC > 0 or logFC < 0). We then identified the optimal k clusters by using the well-established elbow method to identify the smallest ‘k’ that best explained the data, visually described as the elbow (or knee) of a convex plot like that for the sum of squared distances vs k (Fig S4A,C). To do so, we either visually inspected and identified the elbow (Fig 2D) or used the function KneeLocator as implemented in the package Kneed (v 0.7.0) (Fig 4A). We used visual inspection for Fig 2D as we observed that given two k-values (e.g., k = 2,3) with similar sum of squared distances, KneeLocator would choose the larger value whereas we preferred a simpler model. Having identified the optimal number of clusters, ‘k’, we labeled every differentially expressed gene with its assigned cluster and visualized average behavior (median) and the 95% CI (bootstrapped using 1000 iterations) per cluster using Seaborn line plot (v 0.10.0).

To confirm that the identified patterns were not spurious (i.e., an artifact of the k-means clustering algorithm), we permuted the data columns (logFC per timepoint) for each gene thereby scrambling any time-related structure while preserving its overall distribution. We then visualized the result using Seaborn line plot as described above. Following permutation, we observed no longitudinal patterns, which were instead replaced by nearly flat, uninformative trends (Fig S4B,D).

#### Correlation analysis

To verify DEGs identified in the Discovery cohort, we compared logFCs for the Discovery as compared to both Validation 1 and Validation 2 cohorts. We calculated the percentage of genes for which the logFC had the same sign across cohorts (i.e., both positive or both negative) and the spearman correlation using the function scipy.stats.spearmanr. We did not calculate logFCs for DEGs at ≤12 weeks of gestation in Validation 1 because of small sample numbers (3 PE samples prior to 12 weeks).

We also sought to compare whether symptom severity (without or with severe) correlated with logFC magnitude for 544 DEGs identified as common to all PE in design 1. To do so, we calculated the slope of a best-fit line where x and y were defined as logFCs for PE without (x) and with (y) severe features vs NT. We would expect a slope > 1 and < 1 if logFC magnitudes for PE with as compared to without severe symptoms were larger or smaller on average respectively. Similarly, a slope = 1 would reflect that symptom severity did not correlate with logFC magnitude for the 544 DEGs tested.

Finally, to confirm that the identified correlations were significant, we permuted the data columns (logFC per cohort) for each gene thereby scrambling any structure while preserving its overall distribution. We then calculated the same statistics. Following permutation, we observe about 50-55% logFC agreement, as expected at random, a spearman correlation of 0, and slope of 0.

#### Defining cell-type and tissue-specific gene profiles

Cell-type and tissue specific gene profiles were all identified as previously described ^44^. We also briefly summarize this method below.

On the tissue level, for genes and tissues (and some blood and immune cell types) measured in the Human Protein Atlas (HPA, v19)^55^, we calculated a Gini index per gene as a measure of tissue specificity. As a measure of inequality, Gini index values closer to 1 represent genes that are tissue specific. We defined a given gene Y as specific to tissue X if Gini(Y) ≥ 0.6 and max expression for Y is in tissue X. Although the aforementioned method identifies fairly tissue specific genes, it is possible to have a gene Y where Gini(Y) ≥ 0.6 and the gene is expressed in more than 1 tissue (e.g., enrichment in placenta and muscle). To this end, when tracking cell-type and tissue trajectories over gestation (e.g., Fig 4B), where the specificity of a given gene profile is especially important, we imposed a further constraint to ensure that any gene signal only reflects the named tissue (e.g., any gene named placenta specific is specific to the placenta alone). Specifically, we required that genes be annotated by HPA as ‘Tissue enriched’ or ‘Tissue enhanced’ and term this reference, HPA strict.

On the cell-type level, we augmented Tabula Sapiens v1.0 (TSP) with cell types from missing (e.g., placenta, brain), incompletely described tissues (e.g., kidney), or additional annotations (e.g., liver) known to be important in PE. We term this augmented reference TSP+. For genes and cell types measured in Tabula Sapiens v1.0 (TSP), we defined a given gene Y as specific to cell-type X if Gini(Y) ≥ 0.8 and max mean expression for Y is in cell-type X. We combined annotations for all neutrophil and endothelial subtypes as these were based on manifold clustering and it was unclear if the subtypes were truly distinct enough to be distinguished at a whole-body level for our purposes. For genes and cell types described in individual tissue single-cell atlases, we required that a gene be differentially expressed in the specific single cell atlases and tissue specific per the HPA (Gini ≥ 0.6). The following single cell atlases were used for each organ: 1) Placenta ^64,65^ 2) Liver ^66^ 3) Kidney ^67^ 4) Heart ^68^ 5) Brain ^69^.

For TSP+, we then took the union of TSP and individual atlas gene annotations. A small number of genes had conflicting double annotations in TSP as compared to at most one individual tissue single cell atlas. In these rare instances, which most often occurred for genes related to cell-types missing in TSP (e.g., placental or brain cell-types), we preferred the individual single-cell atlas label to TSP.

#### Determining cell type and tissue of origin

We determined whether a given cell-type or tissue was enriched in PE by comparing PE DEGs with cell-type and tissue gene profiles using a hypergeometric test. For every tissue (HPA) or cell-type (TSP+) with at least 2 DEGs specific to it, we performed the following. First, we defined a hypergeometric distribution (scipy.stats.hypergeom, (v 1.5.1)) with the following parameters where category refers to tissue when using HPA and cell-type when using TSP+: M = Number of genes specific to any category, n = Number of genes specific to this category, N = number of DEGs in this k-means longitudinal cluster specific to this category. Next, we estimated a p-value using the survival function (1-CDF) for the specified distribution. Specifically, a p-value is defined as the cumulative probability, prob(X>(n_DEGs_specific_to_this_category–1)), that the distribution takes a value greater than the number of DEGS specific to this category – 1. Finally, once we estimated a p-value for every cell-type (TSP+) or tissue (HPA) identified in each DEG k-means longitudinal cluster, we adjusted for multiple hypotheses using Benjamini-Hochberg and a significance threshold of 0.05.

#### Defining relative signature score per cell type or tissue

We define a signature score as the sum of logCPM values over all genes in a given tissue or cell-type gene profile. We required that a cell type or tissue gene profile have at least 5 specified genes to be considered for signature scoring in cfRNA. Genes were defined as specific to a given tissue based on the reference, HPA strict, and to a given cell type, based on the reference TSP+ (see “Defining cell-type and tissue-specific genes” for details).

To account for our prior observation that baseline cfRNA levels vary per subject – the consequence of biological and technical (e.g., sample processing) factors, we chose to calculate relative as opposed to absolute signature scores. For each subject for which the post-partum sample passed sample QC (see “Sample quality filtering” for details), we estimated a relative signature score defined as the difference between the signature score at a given gestational time point and the post-partum sample. For both Discovery and Validation 1, 49 NT and 24 PE subjects had a post-partum sample that survived sample QC. After normalization, all samples at post-partum had a similar baseline (0). We note that one can define a relative signature score based on any sampled time point for a given person. We chose the post-partum sample because we were interested in tracking maternal organ health over gestation.

Finally, we scaled (i.e., z-score) the relative signature scores for a given cell type or tissue by dividing by the interquartile range, a robust alternative to standard deviation, using the sklearn.preprocessing class, RobustScaler. This accounted for differing gene profile lengths and gene expression levels, and allowed us to compare both different cell-type and tissue contributions and case groups per cell-type or tissue.

Having defined a relative signature score per cell-type and tissue, we visualized average behavior (median) and the 75% CI, a non-parametric estimation of standard deviation, (bootstrapped relative signature score per case group and timepoint using 1000 iterations) using Seaborn line plot (v 0.10.0).

Functional enrichment ***analysis***

Functional enrichment analysis was performed using the tool, GProfiler (v1.0.0) for the following data sources, Gene ontology: biological processes and cellular compartments (GO:BP, GO:CC, released 2021-05-01), Reactome (REAC, released 2021-05-07), and Kyoto Encyclopedia of Genes and Genomes (KEGG, released 2021-05-03). To identify GO terms, we excluded electronic GO annotations (IEA) and used a custom background of only the 7160 genes that were included in DE. We then performed the recommended multiple hypothesis correction (g:SCS) with an experiment wide significance threshold of a = 0.05 ^70^.

#### Logistic regression feature selection and training

To build a robust classifier that can identify mothers at risk of PE at or before 16 weeks of gestation, we first pre-selected features using the set of 330 DEGs when contrasting PE vs NT (see design 2 in “Differential expression analysis” and Supplementary Note 2) as a starting point.

We normalized gene measurements using a series of steps. First, to correct for batch effect, where we define batch as a set of samples processed at the same time by a distinct group (e.g., Discovery cohort = batch, Del Vecchio and colleagues’ cohort = batch), we centered the data by subtracting the median logCPM per gene for a given cohort. Next, we scaled gene values for each cohort using its corresponding interquartile range in the Discovery cohort. Finally, to account for sampling differences across sample, we used an approach similar to when analyzing RT-qPCR data, and normalized data using multiple internal control (i.e., housekeeping) genes. On a per sample basis, we subtracted the median, normalized logCPM value (centered and scaled) for all internal control genes, which we define as 66 genes for which the measured value did not change across PE vs NT comparisons (All genes with adjusted p-value > 0.99 for PE vs NT, Design 2). When calculating the median value for all internal control genes, we excluded any 0 logCPM values as these were likely the consequence of technical dropout.

Model training then used the Discovery cohort alone split into 80% for hyperparameter tuning and 20% for model selection and consisted of two stages – further feature pre-selection based on two metrics followed by the construction of a logistic regression model with an elastic net penalty. Using a split Discovery cohort for training mitigated overfitting even though all Discovery samples were used for differential expression, which defined the initial feature set.

For feature pre-selection, we calculated logFC values using the 80% Discovery split for all 330 genes for PE vs NT. We focused on two practical metrics measured across the 80% split of Discovery samples collected on or before 16 weeks of gestation: gene change size (|logFC|) and gene change stability (CV). All model hyperparameters were then tuned using AUROC as the outcome metric and 5-fold cross validation. Next, we selected the best model including tuned feature pre-selection cutoffs again using AUROC. Specifically, we calculated an AUROC score for both the 80% and 20% Discovery splits separately, and the selected model achieved the best score on both splits

Finally, we tuned the probability threshold, P, at which a sample is labeled as at risk of PE if prob(PE) ≥ P using the entire Discovery cohort. To do so, we constructed a receiver operator characteristic curve (ROC) and calculated the false positive rate (FPR) and true positive rate (TPR) at different thresholds, P_i,. We identified the threshold, P_i, at which FPR=10%, and round to the nearest 5 (e.g., 0.37 would become 0.35). This yielded a tuned threshold of P = 0.35. All classifications as negative or positive were then made based on this threshold.

To understand the importance of each gene feature, we trained a separate logistic regression model for a subset of all possible feature subset (307 combinations out of a total of 262143 for 1–17 genes). No feature pre-selection was performed for this sub-analysis. All model hyperparameters were tuned as previously described. We defined a gene subset as weakly predictive if the model yielded an AUROC > 0.5 on the test set (Validation 2).

In all cases, performance metrics were assessed as described below (see next section) and used Scikit-learn (v 0.23.2),

#### Performance metric analysis

Model performance was assessed using several statistics including sensitivity, specificity, PPV, NPV, and AUROC. Given a 2×2 confusion matrix where rows 1and 2 represent true negatives and positives and columns 1 and 2 represent negative and positive predictions respectively, we can define the value in row 1, column 1 as true negatives (TN), row 1, column 2 as false positives (FP), row 2, column 1 as false negatives (FN), and row 2, column 2 as true positives (TP). We can then define the following proportions: (1) Sensitivity = TP / (TP + FN) (2) Specificity = TN / (TN + FP) (3) PPV = TP / (TP + FP) (4) NPV = TN / (TN + FN). For each proportion, we calculated 90% CIs using Jeffrey’s interval ^71^ and the function, proportion_confint, from statsmodels.stats.proportion. We also approximated AUC and its corresponding 90% CI using the Scikit-learn function, roc_auc_score, and the binormal approximation respectively.

### Statistical analyses

All p-values reported herein were calculated using the non-parametric Mann-Whitney rank test unless otherwise stated. One-sided tests were performed where required based on the hypothesis tested.

### Code/data availability

All computational analyses were performed using Python 3.6 or R 3.5, and will be available on Github. Raw and processed sequencing data will be deposited with the SRA and GEO, respectively.

## Supplementary text

### Supplementary note 1: Establishing quality metrics to identify sample outliers

Because cfRNA measurements can be noisy ^31,72^, we have previously developed and reported on three quality metrics that can flag sequenced cfRNA samples with poor quality ^62,63^. Specifically, these metrics aim to quantify unusually high levels of RNA degradation and/or DNA contamination by comparing a given sample’s value for any of these metrics with what we expect empirically. We defined reasonable expected values for each metric based on the 95^th^ percentile for ∼700 previously sequenced samples across 3 cohorts.

We found that samples with outlier values for at least one of these metrics both clustered separately and served as leverage points in PCA (Fig S1A-C). To avoid introducing unwanted bias, we removed these low-quality samples from any further analysis. After removing outlier samples, we reran PCA and noticed that some samples continued to serve as leverage points. We suspected that this may be due to genes that were poorly detected and consequently performed further filtering to identify well-detected genes across the entire cohort. Specifically, we used a basic cutoff that required a given gene be detected at a level of at least 0.5 CPM reads in at least 75% of samples after removing outlier samples. Following this step, we retain 7,160 genes for analysis. Upon inspection, we find that visualization using PCA is no longer driven by leverage points.

### Supplementary note 2: Selecting an initial feature set for machine learning

We first explored whether a common gene set could describe PE with or without severe features. We observed that we could separate PE from NT samples (Fig 2) irrespective of symptom severity and that PE with or without severe features as compared to NT had on average the same log FC (Fig S2E). With this in mind, we reran differential expression to identify a core set of genes that can distinguish PE (as a binary case group) from NT (See Design 2 in methods section “Differential expression analysis” for more details). This identified 330 genes that we used as an initial feature set for machine learning.

**Fig S1.**
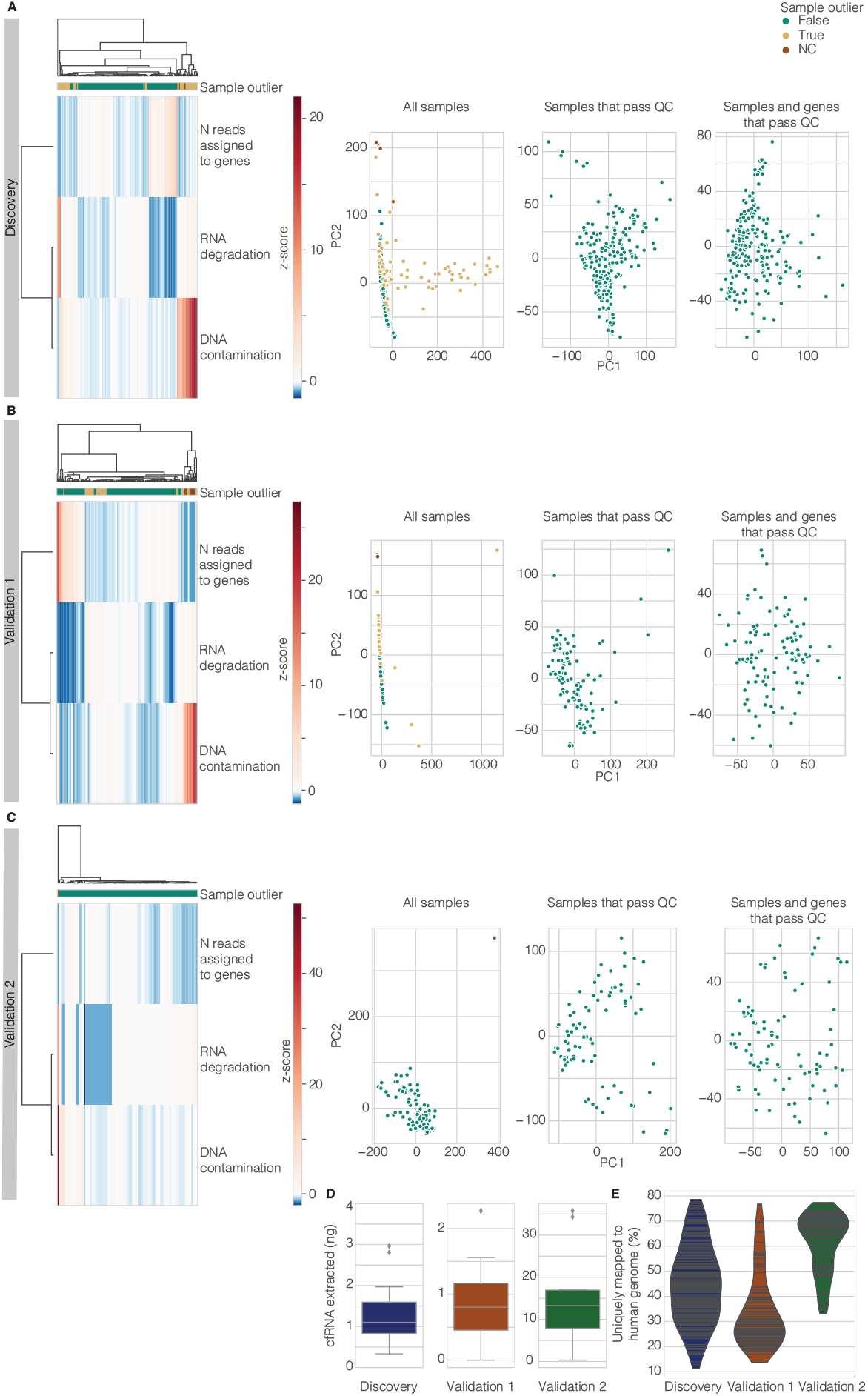
Samples with outlier values for at least one of QC metric cluster separately from most non-outlier samples. For Discovery (**A**), Validation 1 (**B**), and Validation 2 (**C**), hierarchical clustering (left) and PCA reveals that most outlier samples cluster with negative control (NC) samples (H2O) and separately from non-outlier samples. (**D, E**) Visualization of other QC metrics like the amount of cfRNA extracted (**D**) and the percent of reads that align uniquely to the human genome (**E**). For PCA in (**A-C**), sample outliers and poorly detected genes drive PCA and serve as leverage points. The top two principal components are visualized when performed using all samples and all genes (leftmost PCA) or only samples that pass QC metrics (middle PCA) reveals that certain samples can act as leverage points. Once sample outliers and lowly detected genes are removed from the cfRNA gene matrix (rightmost PCA), the top two principal components reflect natural variance in the data and are no longer driven by a few leverage points.

**Fig S2.**
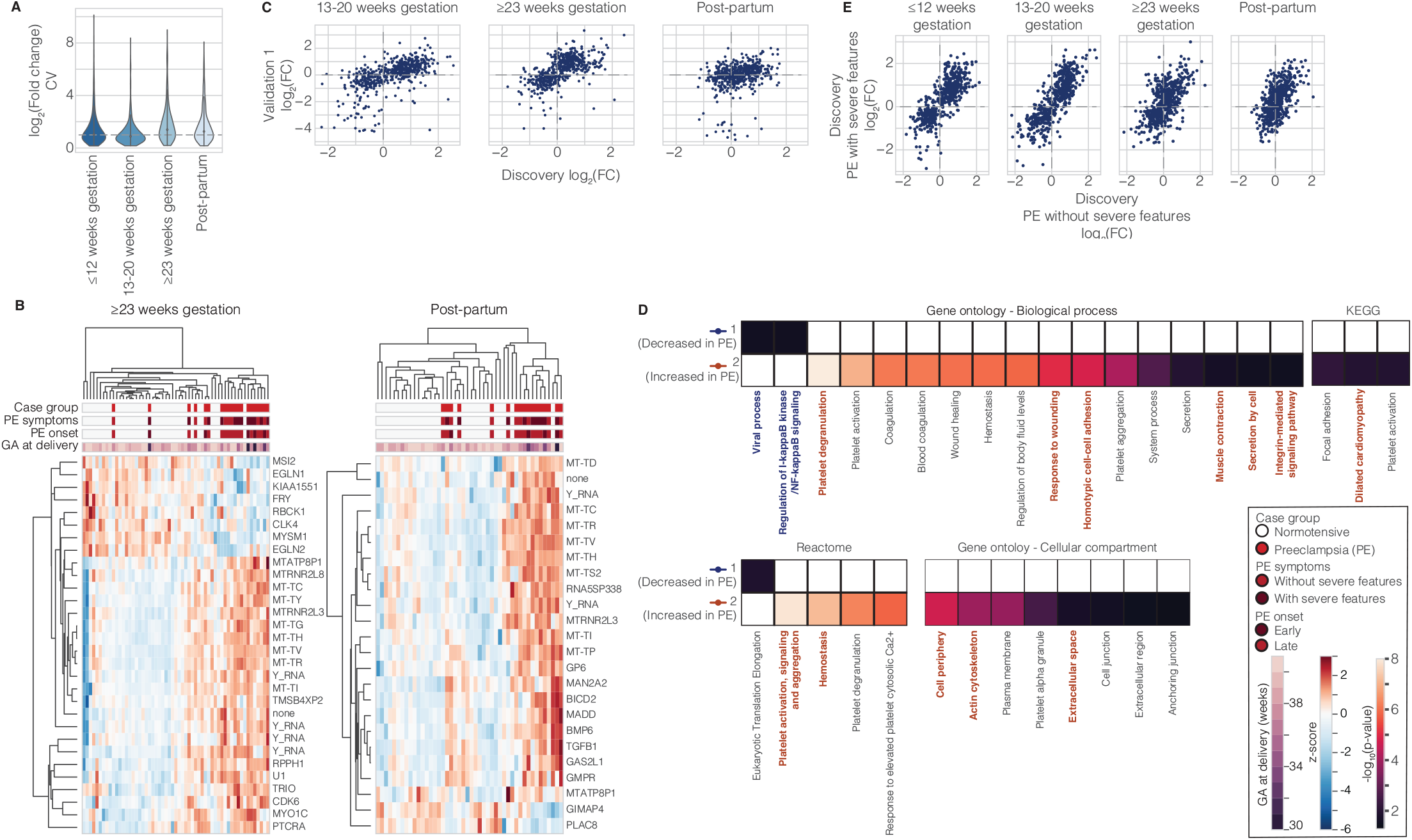
Across gestation prior to diagnosis, changes in the cfRNA transcriptome segregate PE and NT samples and reflect known PE biology. (**A**) Distribution of CVs with dashed line at CV = 1 for all DEGs between PE as compared to NT samples across gestation. (**B**) At ≥23 weeks of gestation and post-partum, in each sample collection period, a subset of DEGs can separate PE and NT samples despite differences in symptom severity, PE onset subtype, and gestational age (GA) at delivery. (**C**) Comparison of log(Fold change) for DEGs for PE as compared to NT between Discovery (x-axis) and Validation 1 (y-axis) reveals good agreement across gestation but not post-partum: 82%, 83%, and 60% of genes had the same logFC sign with a Spearman correlation of 0.67, 0.69, and 0.35 (p < 10^−15^) at 13–20, ≥23 weeks, and post-partum, respectively. **(D)** The genes in each longitudinal trend group reflect known PE etiology as highlighted across four databases (GO biological processes, KEGG, the reactome, and GO cellular compartment). Some PE associated terms are emphasized in bold, colored text that corresponds to group color from Fig 2D (Dark blue and orange indicate decreased and increased in PE vs NT, respectively). **(E)** Comparison of log(Fold change) for DEGs for PE without severe features vs NT (x-axis) and PE with severe features vs NT in the Discovery cohort (y-axis) reveals good agreement along the y=x axis with a slope of 0.93, 1.03, 0.77, and 0.86 at ≤12 weeks, 13–20, ≥23 weeks, and post-partum, respectively.

**Fig S3.**
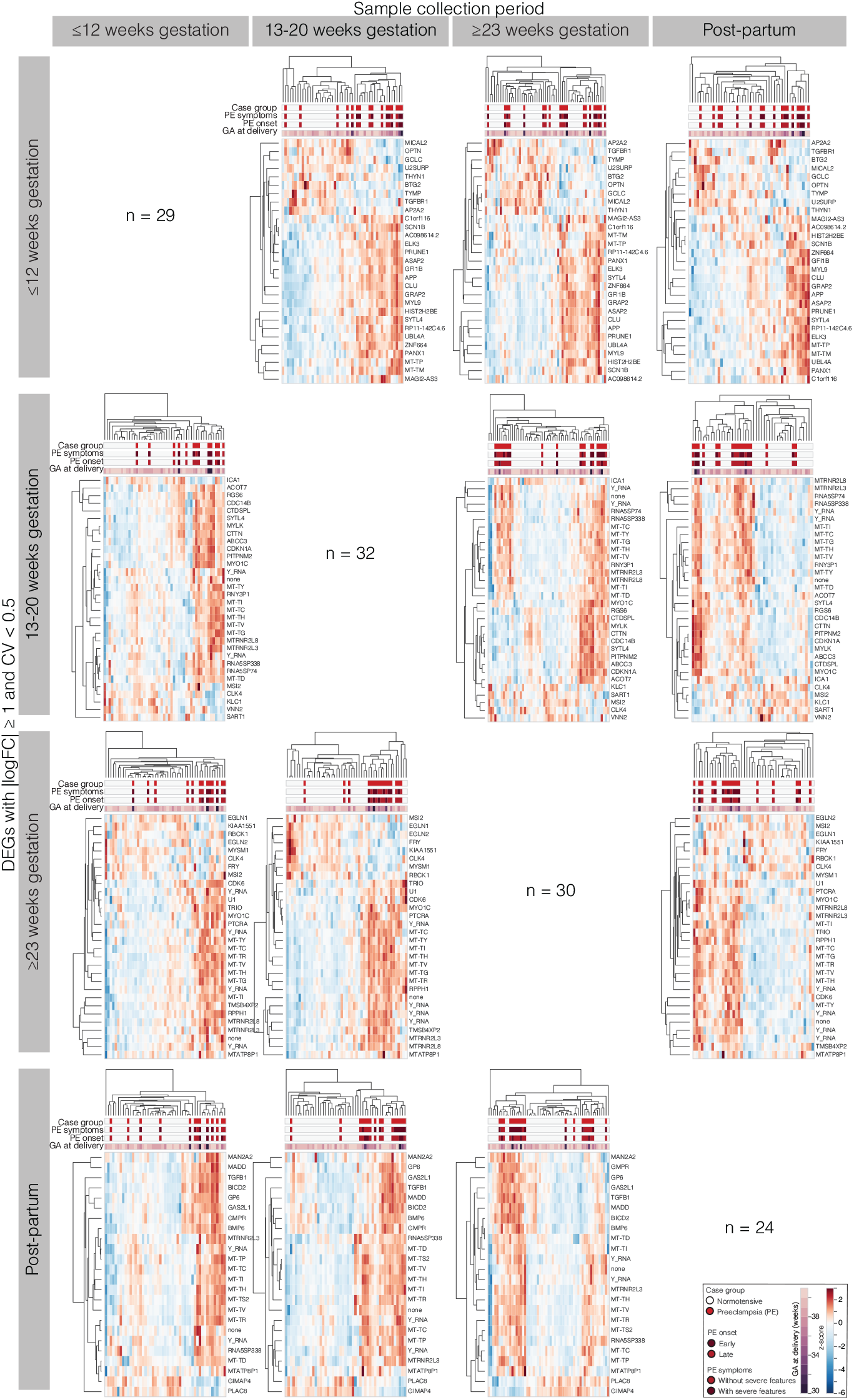
Across gestation and prior to diagnosis, changes in the cfRNA transcriptome identified at one timepoint can moderately segregate PE and NT samples at other timepoints. DEGs with |logFC| ≥ 1 and CV < 0.5 or 0.4 for 13–20 weeks timepoint were identified at each timepoint across gestation. Each row visualizes how well a specific DEG subset from a given sample collection period can separate PE and NT samples in all other collection periods (columns). The number of genes identified per sample collection period is highlighted along the main diagonal.

**Fig S4.**
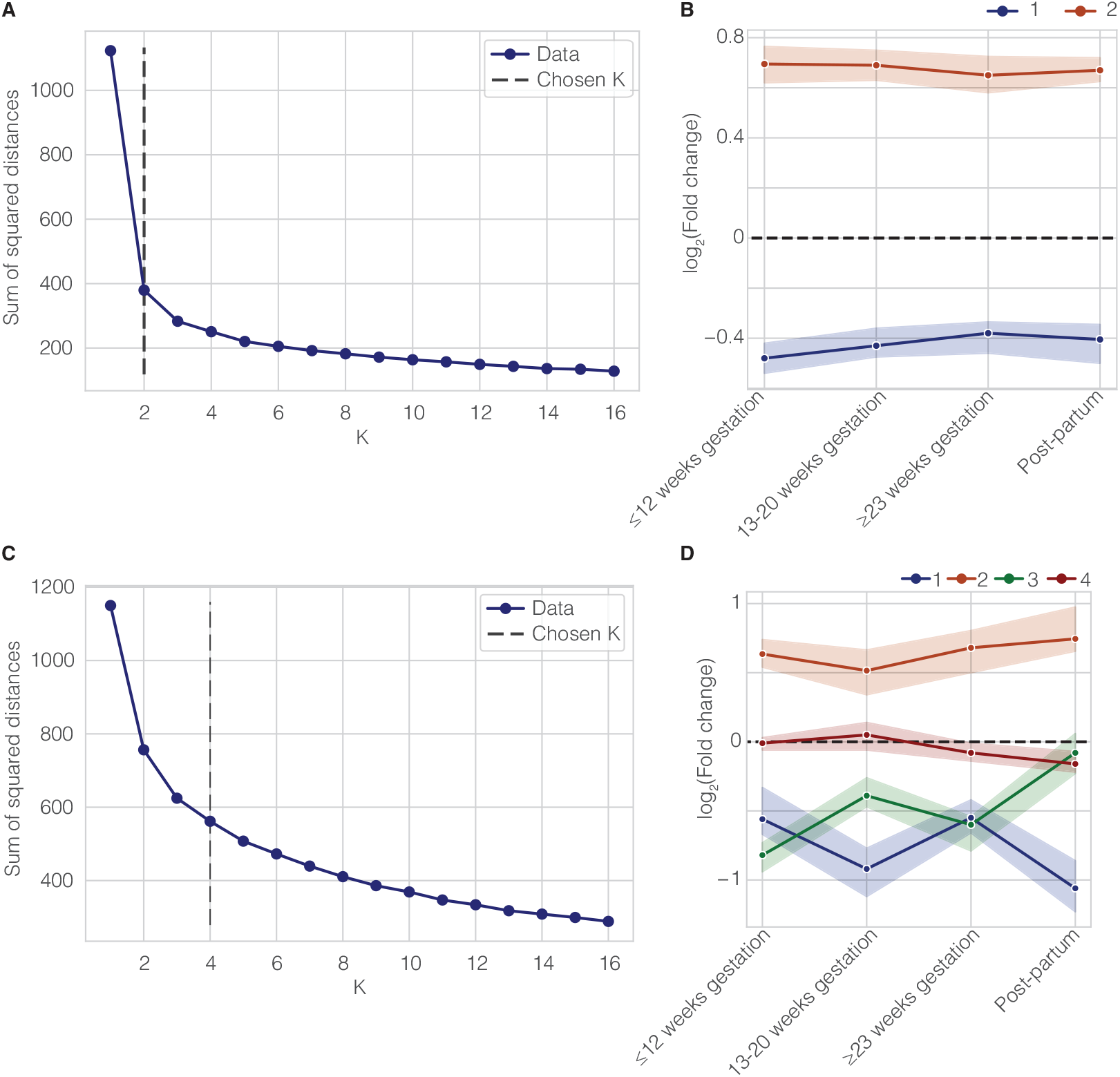
K-means clustering reveals meaningful longitudinal patterns. The chosen k clusters (dashed line) comparing a performance metric, the sum of squared distances, and values of k for clustering of DEGs for PE vs NT related to Fig 2D in (**A**) and DEGsfor PE with vs without severe features related to Fig 4A in (**C**). Following permutation of the data matrix prior to k-means clustering, longitudinal changes over gestation are replaced by (**B**) 2 flat lines for clustering of logFC for PE vs NT and (**D**) 4 uninformative lines for clustering of logFC for PE with vs without severe features.

**Fig S5.**
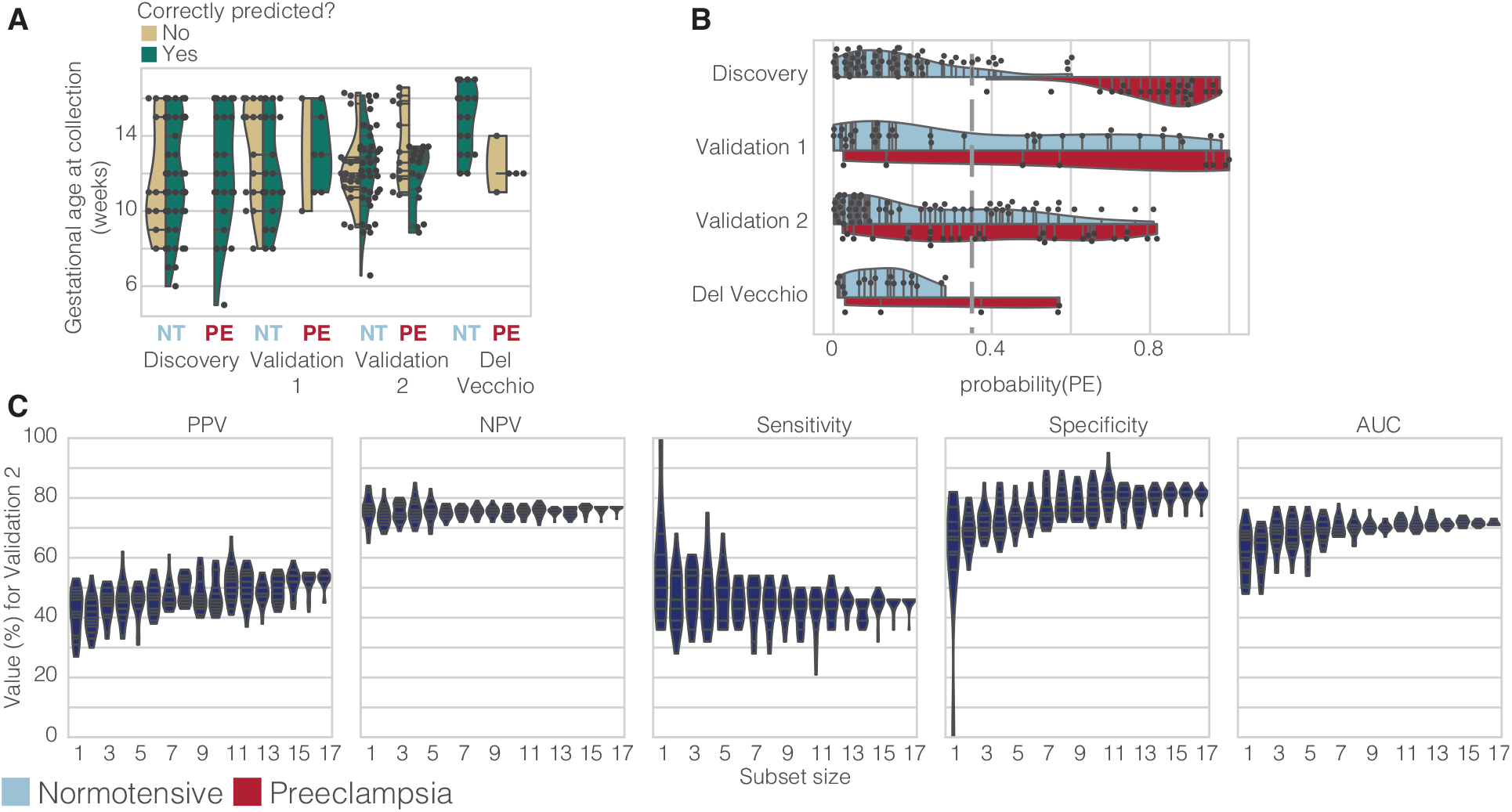
Examining the logisitic regression model used to predict risk of PE early in gestation. (**A**) Comparison of gestational age at sample collection (weeks) for incorrectly predicted (yellow) or correctly predicted (green) samples across NT and PE groups in Discovery, Validation 1, Validation 2, and Del Vecchio shows that incorrectly predicted PE samples (false negatives) are collected at later gestational ages. (**B**) Estimated probability of PE as outputted by logistic regression for both PE and NT samples shows that the model is well-calibrated across most predictions. Dashed line at 0.35 indicates classifier cutoff where probability(PE) ≥ 0.35 constitutes a sample predicted as PE. (**C**) Logistic regression models trained on subsets of 1–18 genes of the initial 18 genes can moderately predict future PE onset in Validation 2 cohort with improving performance as subset size increases and as characterized by PPV, NPV, sensitivity, specificity, and AUROC (left to right).

**Table S1.** Participant, pregnancy, and PE characteristics across both Discovery and Validation cohorts. Maternal age and BMI, gestational age (GA) at delivery, fetal weight, and GA at PE onset are reported mean ± SD. All other values are reported as percentages with the corresponding count in parentheses. Small for GA (SGA) was defined as an infant with a birthweight below the 10^th^ centile for their GA at delivery. Pre-pregnancy BMI was not available for individuals in Validation 2 cohort. AI/AN indicates American Indians and Alaska Natives. *adjusted p ≤ 0.05, chi-squared (categorical) or ANOVA (continuous) test comparing all cohorts ‡ adjusted p ≤ 0.05, chi-squared (categorical) or ANOVA (continuous) test comparing PE and NT within each cohort † denotes that missing values were omitted from reported values for a given feature.

| Maternal pre-pregnancy characteristics |  |  |  |  |  |  |  |  |
| --- | --- | --- | --- | --- | --- | --- | --- | --- |
|  |  | Age, years | BMI, kg/m<sup>2</sup> | Nulliparous, % (n) * | Smoker, % (n) | History of preterm birth, % (n) * | History of PE, % (n) |  |
| Discovery | NT | 32 ± 5 | 23 ± 3 †,‡ | 31 (15) | 2 (1) | 16 (8) | 2 (1) |  |
|  | PE | 31 ± 6 | 30 ± 8 ‡ | 38 (9) | 0 (0) | 25 (6) | 17 (4) |  |
| Validation 1 | NT | 31 ± 5 | 25 ± 6 † | 28 (9) | 3 (1) | 12 (4) | 9 (3) |  |
|  | PE | 34 ± 5 | 26 ± 4 | 29 (2) | 0 (0) | 29 (2) | 29 (2) |  |
| Validation 2 | NT | 30 ± 5 | NA | 0 (0) | 5 (3) | 0 (0) ‡ | 0 (0) |  |
|  | PE | 32 ± 6 | NA | 0 (0) | 0 (0) | 50 (13) ‡ | 12 (3) |  |
| Maternal ethnicity/race |  |  |  |  |  |  |  |  |
|  |  | Ethnicity * | Race, % (n) * |  |  |  |  |  |
|  |  | Hispanic | White | AI/AN | Black | Asian | Other | Unknown |
| Discovery | NT | 22 (11) | 73 (36) | 0 (0) | 4 (2) | 20 (10) | 2 (1) | 0 (0) |
|  | PE | 42 (10) | 50 (12) | 0 (0) | 4 (1) | 25 (6) | 0 (0) | 21 (5) |
| Validation 1 | NT | 34 (11) | 72 (23) | 0 (0) | 6 (2) | 16 (5) | 6 (2) | 0 (0) |
|  | PE | 57 (4) | 71 (5) | 0 (0) | 0 (0) | 14 (1) | 0 (0) | 14 (1) |
| Validation 2 | NT | 8 (5) | 59 (36) | 23 (14) | 5 (3) | 8 (5) | 0 (0) | 5 (3) |
|  | PE | 4 (1) | 62 (16) | 23 (6) | 4 (1) | 4 (1) | 0 (0) | 4 (1) |
| Pregnancy characteristics |  |  |  |  |  |  |  |  |
|  |  | GA at delivery, weeks * | Mode of delivery % vaginal (n) * | Multi-gestation, % (n) | Preterm delivery, % (n) * | Fetal sex, % male (n) | Fetal weight, kg * | SGA, % (n) |
| Discovery | NT | 39 ± 1 ‡ | 76 (37) | 0 (0) | 0 ‡ | 53 (26) | 3.4 ± 0.4 ‡ | 0 (0) |
|  | PE | 36 ± 3 ‡ | 46 (11) | 12 (3) | 38 (9) ‡ | 54 (13) | 2.8 ± 0.7 ‡ | 4 (1) |
| Validation 1 | NT | 39 ± 1 | 56 (18) | 3 (1) | 0 (0) | 53 (17) | 3.4 ± 0.5 † | 0 (0) |
|  | PE | 38 ± 1 | 57 (4) | 0 (0) | 14 (1) | 71 (5) | 3.4 ± 0.3 | 0 (0) |
| Validation 2 | NT | 39 ± 1 ‡ | 75 (46) ‡ | 0 (0) | 0 (0) ‡ | 46 (28) | 3.4 ± 0.4 ‡ | 0 (0) |
|  | PE | 37 ± 2 ‡ | 35 (9) ‡ | 8 (2) | 42 (11) ‡ | 54 (14) | 2.9 ± 0.7 ‡ | 8 (2) |
| PE characteristics |  |  |  |  |  |  |  |  |
|  |  | GA at onset, weeks |  | Onset type, % early (n) |  | Symptom severity, % severe (n) |  |  |
| Discovery | PE | 36 ± 4 |  | 25 (6) |  | 54 (13) |  |  |
| Validation 1 | PE | 38 ± 2 |  | 0 (0) |  | 43 (3) |  |  |
| Validation 2 | PE | 38 ± 2 † |  | 12 (3) † |  | 50 (13) † |  |  |

**Table S2.** Number of subjects with a given number of samples that passed QC for Discovery and Validation 1 cohorts.

|  | Number of samples |  |  |  |  |
| --- | --- | --- | --- | --- | --- |
|  | 1 | 2 | 3 | 4 | 5 |
| **Discovery** | 5 | 19 | 31 | 17 | 1 |
| **Validation 1** | 7 | 8 | 14 | 9 | 1 |

**Table S3.**
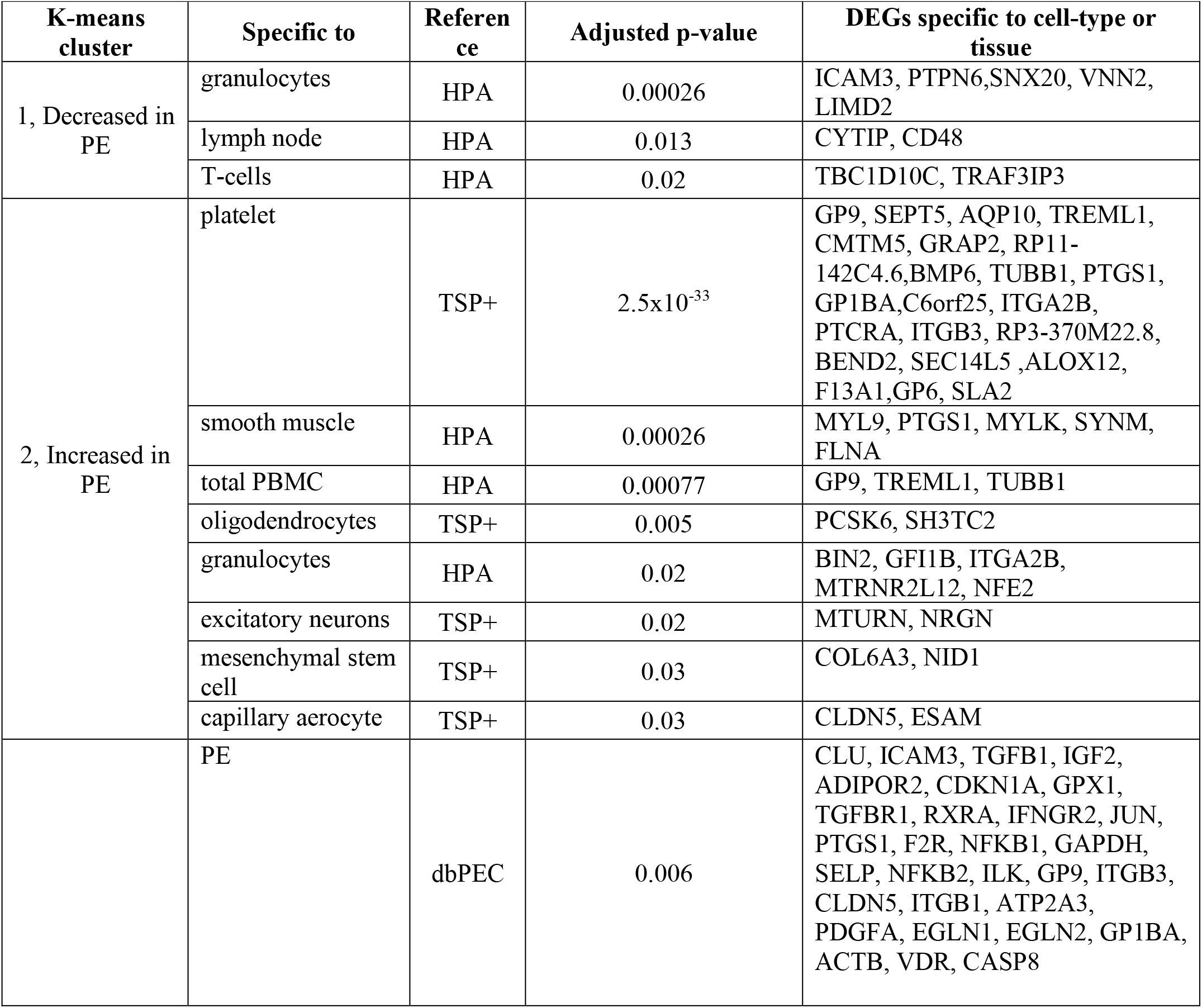
Tissue, cell-types, and genes previously implicated in PE enriched in 544 DEGs identified when comparing PE with or without severe features and NT pregnancies. For every significantly enriched tissue or cell-type (adjusted p ≤ 0.05, Hypergeometric test with Benjamini-Hochberg correction), assigned k-means cluster (i.e., Fig 2D), reference, adjusted p-values, are reported from left to right. Finally, the rightmost column lists the gene names for all DEGs that were labeled as specific to a given cell-type or tissue. The last row lists gene set enrichment with dbPEC, a PE specific database of genes.

**Table S4.** PE prediction relies on the cfRNA levels of 18 genes. For every gene, symbol, ENSEMBL ID, full name, odds ratio (OR) based on the logistic regression coefficient, and if available, a subset of GO biological processes and molecular functions (if available) are reported from left to right.

| Gene | ENSEMBL | Name | Odds ratio | Biological process [GO] | Molecular function [GO] |
| --- | --- | --- | --- | --- | --- |
| CAMK2G | ENSG00000148660 | calcium/calmodulin dependent protein kinase II gamma | 0.65 | nervous system development, protein phosphorylation, regulation of neuron projection development, regulation of skeletal muscle adaptation, insulin secretion, cell differentiation | calcium-dependent protein serine/threonine phosphatase activity, identical protein binding, protein homodimerization activity, |
| DERA | ENSG00000023697 | deoxyribose-phosphate aldolase | 1.07 | pentose-phosphate shunt, deoxyribose phosphate catabolic process | deoxyribose-phosphate aldolase activity, protein binding |
| FAM46A | ENSG00000112773 | terminal nucleotidyl transferase 5A | 0.91 | mRNA stabilization, response to bacterium | RNA adenylyltransferase activity, protein binding, RNA binding |
| KIAA1109 | ENSG00000138688 | KIAA1109 | 0.89 | synaptic vesicle endocytosis, regulation of epithelial cell differentiation, endosomal transport, | protein binding |
| LRRC58 | ENSG00000163428 | leucine rich repeat containing 58 | 0.79 |  |  |
| MYLIP | ENSG00000007944 | myosin regulatory light chain interacting protein | 0.94 | nervous system development, regulation of low-density lipoprotein particle receptor catabolic process, ubiquitin-dependent protein catabolic process, negative regulation of neuron projection development | ubiquitin protein ligase activity, cytoskeletal protein binding, protein binding, metal ion binding, ubiquitin-protein transferase activity |
| NDUFV3 | ENSG00000160194 | NADH:ubiquinone oxidoreductase subunit V3 | 0.66 | mitochondrial electron transport, NADH to ubiquinone | NADH dehydrogenase (ubiquinone) activity, protein binding, RNA binding |
| NMRK1 | ENSG00000106733 | nicotinamide riboside kinase 1 | 1.42 | NAD metabolic process, phosphorylation, NAD biosynthetic process | ribosylnicotinamide kinase activity |
| PI4KA | ENSG00000241973 | phosphatidylinositol 4-kinase alpha | 0.9 | multi-organism membrane organization, phosphorylation, viral replication complex formation and maintenance, signal transduction | kinase activity, phosphatidylinositol kinase activity, protein binding |
| PRTFDC1 | ENSG00000099256 | phosphoribosyl transferase domain containing 1 | 1.32 | guanine salvage, GMP catabolic process, purine ribonucleoside salvage | protein homodimerization activity, hypoxanthine phosphoribosyltransferase activity |
| PYGO2 | ENSG00000163348 | pygopus family PHD finger 2 | 0.95 | positive regulation of chromatin binding, developmental growth, lens development in camera-type eye, regulation of histone H3-K4 methylation, mammary gland development | chromatin binding, protein binding, metal ion binding, histone acetyltransferase regulator activity, histone binding |
| RNF149 | ENSG00000163162 | ring finger protein 149 | 0.82 | ubiquitin-dependent protein catabolic process, regulation of protein stability, protein ubiquitination, | ubiquitin protein ligase activity, metal ion binding |
| TFIP11 | ENSG00000100109 | tuftelin interacting protein 11 | 0.9 | biomineral tissue development, negative regulation of protein binding, spliceosomal complex disassembly, mRNA splicing, via spliceosome, | protein binding, nucleic acid binding |
| TRIM21 | ENSG00000132109 | tripartite motif containing 21 | 0.88 | negative regulation of protein deubiquitination, regulation of protein localization, regulation of gene expression, protein monoubiquitination, | zinc ion binding, transcription coactivator activity, identical protein binding |
| USB1 | ENSG00000103005 | U6 snRNA biogenesis phosphodiesterase 1 | 0.85 | U6 snRNA 3'-end processing, RNA splicing, | poly(U)-specific exoribonuclease activity |
| Y_RNA | ENSG00000201412 | Y RNA | 1.85 |  |  |
| Y_RNA | ENSG00000238912 | Y RNA | 1.08 |  |  |
| YWHAQ P5 | ENSG00000236564 | YWHAQ pseudogene 5 | 1.09 |  |  |

**Table S5.**
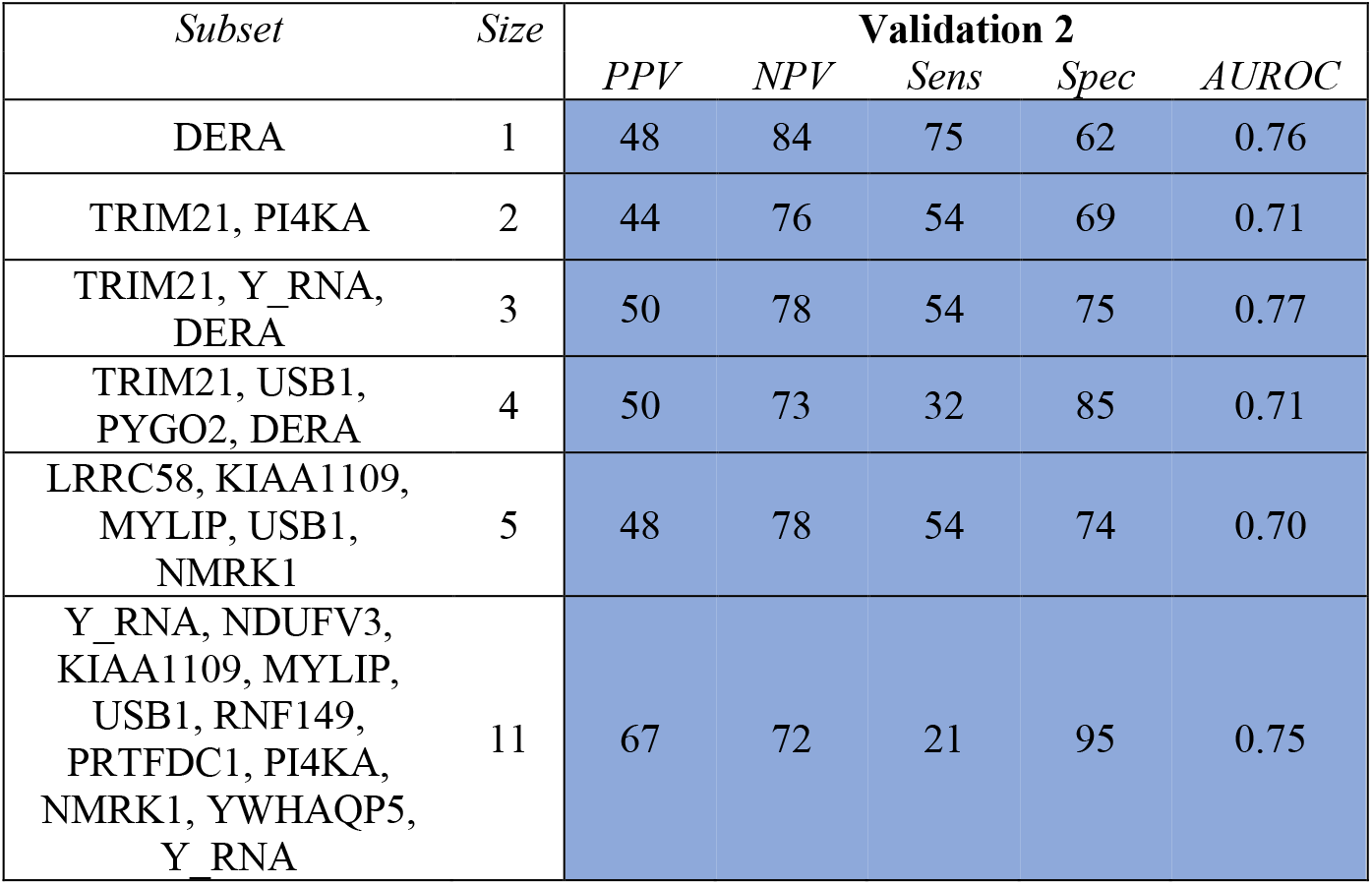
Logistic regression models trained on some subsets of 1–18 genes of the initial 18 genes can predict future PE onset with nearly equivalent performance metrics. The associated performance metrics for each data split and some high-performing gene subsets is reported including sensitivity (Sens), specificity (Spec), PPV, NPV, and AUROC, which are reported as the estimated percentage. Only a few, illustrative examples are shown here.

| *Subset* | *Size* | **Validation 2** |  |  |  |  |
| --- | --- | --- | --- | --- | --- | --- |
|  |  | *PPV* | *NPV* | *Sens* | *Spec* | *AUROC* |
| DERA | 1 | 48 | 84 | 75 | 62 | 0.76 |
| TRIM21, PI4KA | 2 | 44 | 76 | 54 | 69 | 0.71 |
| TRIM21, Y<sub>RNA</sub>, DERA | 3 | 50 | 78 | 54 | 75 | 0.77 |
| TRIM21, USB1, PYGO2, DERA | 4 | 50 | 73 | 32 | 85 | 0.71 |
| LRRC58, KIAA1109, MYLIP, USB1, NMRK1 | 5 | 48 | 78 | 54 | 74 | 0.70 |
| Y<sub>RNA</sub>, NDUFV3, KIAA1109, MYLIP, USB1, RNF149, PRTFDC1, PI4KA, NMRK1, YWHAQP5, Y<sub>RNA</sub> | 11 | 67 | 72 | 21 | 95 | 0.75 |

**Table S6.** Tissue and cell-types enriched in 503 DEGs identified when comparing PE with as compared to without severe features. For every significantly enriched tissue or cell-type (adjusted p ≤ 0.05, Hypergeometric test with Benjamini-Hochberg correction), assigned k-means cluster (i.e., Fig 4A), reference, adjusted p-values, are reported from left to right. Finally, the rightmost column lists the gene names for all DEGs that were labeled as specific to a given cell-type or tissue.

| K-means cluster | Specific to | Reference | Adjusted p-value | DEGs specific to cell-type or tissue |
| --- | --- | --- | --- | --- |
| 1, Increased across gestation for severe PE | naive b cell | TSP+ | 0.0002 | TCL1A,TNFRSF13C |
|  | B-cells | HPA | 0.002 | FCMR,TCL1A |
|  | neutrophil | TSP+ | 0.002 | PADI4,MEFV,DOK3 |
|  | bone marrow | HPA | 0.01 | PADI4,DOK3 |
|  | endothelial cell | TSP+ | 0.04 | PDE2A,VWF |
| 2, Increased and then decreased in early and late gestation respectively for severe PE | granulocytes | HPA | 0.0007 | SELL,HK3,MXD1,ALPL |
|  | neutrophil | TSP+ | 0.02 | FCN1,HK3 |
| 4, Decreased across gestation for severe PE | thymus | HPA | 0.01 | CARMIL2,ARHGAP15 |

**File S1. Table of 89 differentially expressed genes included in Fig 2B, S2B heatmaps**. For every gene, the symbol, ENSEMBL ID, sample collection groups for which the gene passed cutoff thresholds, full name, ENSEMBL gene type, and a subset of GO biological processes and molecular functions are reported.

**File S2. Table of all significant GO terms related to 544 DEGS for PE with or without severe features as compared with NT**.

